# Multidomain Therapy for Alzheimer’s Disease: A Scoping Review

**DOI:** 10.1101/2025.06.15.25329293

**Authors:** Jared C. Roach, Gwênlyn Glusman, Molly K. Rapozo, David A. Merrill, Jennifer Bramen, John F. Hodes, Prabha Siddarth, Somayeh Meysami, Shannel H. K. Elhelou, Ryan M. Glatt, Lance Edens, Cory Funk, Dan Kelly, William R. Shankle, Dale Bredesen, Cyrus A. Raji, Leroy Hood

**Affiliations:** Institute for Systems Biology, Seattle, WA, USA; Pacific Neuroscience Institute, Santa Monica, CA, USA; Department of Translational Neurosciences, Saint John’s Cancer Institute, Santa Monica, CA, USA; Pickup Family Neurosciences Institute, Hoag Hospital; Department of Neurobiology and Behavior, University of California, Irvine; Shankle Clinic, Newport Beach, CA; EMBIC Corporation, Newport Beach, CA, USA; Departments of Radiology and Neurology, Washington University in St. Louis, St. Louis, MO, USA

**Keywords:** diet, exercise, anti-amyloid, cognitive training, dementia, cognitive decline, Alzheimer’s disease and related disorders (ADRD), Alzheimer’s disease (AD), personalized coaching, systems biology, multimodal interventions, multidomain, multidimensional, multiomics, multi-omics, cognitive impairment, lifestyle

## Abstract

**Background:** Alzheimer’s disease (AD) leading to cognitive decline and dementia results from the interplay of multiple interacting dysfunctional biological systems. These systems can be categorized by <u>domain</u>, such as inflammation, cardiovascular health, proteostasis, or metabolism. Specific causes of AD differ between individuals, but each individual is likely to have causes stemming from multiple domains. Personalized multidomain therapy has been proposed as a standard of care for AD.

**Objectives:** We sought to enumerate and describe prospective randomized controlled trials (RCTs) for multidomain interventions for AD, and to extract their inclusion criteria, trial design parameters (length, number of participants), and outcome measures. We sought to clarify gaps and opportunities in research and clinical translation.

**Eligibility criteria:** We include all cohort studies and RCTs for multidomain (also known as multimodal, multicomponent, multidimensional, or multisystem) therapy of any stage of AD.

**Results:** There have been 22 studies (completed or reported as ongoing) of multidomain interventions for AD, including 18 RCTs. Of the 14 completed RCTs, 11 demonstrate benefit from their intervention in at least one arm.

**Conclusions:** Although these RCTs differ widely in their parameters, the majority support the use of multidomain therapy, and show effect sizes greater than reported for unimodal therapies, including pharmaceuticals. Multidomain therapy should be the standard of care for AD. Multidomain interventions (also known as treatments) should be employed widely, early, and first-line. Treatment or prevention is likely to be most effective at early, presymptomatic stages, but is worthwhile at all stages of disease. In order to influence multiple domains, multiple modes of therapy are likely necessary in all patients. Some individual modes, such as particular lifestyle interventions, may target multiple domains. Nevertheless, most patients will benefit from multiple modes of intervention (multimodal intervention) that together target multiple domains. Standard-of-care guidelines should explicitly include multidomain interventions. Future clinical trials must be designed to iteratively improve multidomain therapies. Payors should embrace reimbursement for effective multidomain intervention, including personalized coaching.

## INTRODUCTION

Alzheimer’s disease (AD) results from the interplay of multiple interacting dysfunctional biological systems, or “domains”—such as inflammation, proteostasis, or metabolism. The causes of AD may stem from multiple genetic, immunological, and environmental factors. As such, AD is considered a “complex disease”.

Translational research has enabled remarkable progress over the last fifty years in developing and deploying therapies for complex diseases such as hypertension and type 2 diabetes. Complex diseases—including AD— typically require complex therapies to prevent and treat dysfunction across many biopsychosocial domains and scales.^1^ Each of these domains of dysfunction can be treated by one or more interventions (also known as treatments) targeting that domain (Sikkes et al, 2021). Specific causes of AD differ between individuals. Personalized, multidomain therapies are needed to best prevent and treat AD.

Multidomain interventions can slow progression, ameliorate, halt, or even reverse the course of complex diseases. In this Perspective, we discuss real-world approaches for applying existing knowledge to clinical care, maximizing knowledge from clinical trials and coupling that research to actionable interventions. We focus on the design and implementation of multidomain interventions, how to deploy these in the real world, and how to learn from these using dynamic dense-data clinical trials. We emphasize that lifestyle interventions, such as exercise and diet, target multiple domains; current evidence suggests they are the most effective treatment for AD (Sikkes et al., 2021). We highlight remote and/or AI coaching as an approach for delivering complex interventions.

### Alzheimer’s disease and Alzheimer’s dementia

A divergence between the definitions of Alzheimer’s disease (AD) and Alzheimer’s dementia began to emerge in the late 20th century and became more pronounced in the early 21st century (George et al., 2013; Jagust, 2021). Alzheimer’s dementia is now defined as a very late stage of AD; these two “AD” acronyms may be conflated in some sources. Few authorities currently offer precise definitions of AD. Such definitions need to be useful both for researchers and for clinicians. It is difficult to come up with a definition suitable for both. Researchers generally desire narrow precise definitions that enable specific hypothesis testing and comparability between studies. Clinicians tend to favor broader definitions that enable diagnosis and treatment of patients in modern medical systems. However, there is some consensus bridging more clinical and more academic stakeholders: AD involves both cognitive decline and measurable pathology with alterations in AD-related molecular subsystems. AD was originally defined primarily as a clinicopathological entity in which the clinical symptoms of dementia were linked to specific amyloid plaque and neurofibrillary tangle neuropathological findings at autopsy (Knopman et al., 2019). However, over the first several decades of the twenty-first century, neuroimaging techniques and liquid (blood and cerebrospinal fluid) biomarkers have been increasingly successful at separating cognitive decline due to AD from other causes. In some cases, the molecular pathology of AD can be identified in individuals without any signs of cognitive decline (Ross & Dodel, 2025). The recent definition of AD published by the Alzheimer’s Association (AA) comes close to fulfilling the need for a precise research definition (Jack et al., 2024). However, exact thresholds and recommendations or standardizations of biomarkers continue to evolve. A precise definition of AD remains elusive. Furthermore, the AA definition may neither fulfill the needs of clinicians nor capture the importance of tau pathology (Dubois et al., 2024; Petersen et al., 2024).

Understanding the mechanistic or causal links between AD biomarkers, pathology, and symptoms is advancing steadily. Epistemic theories of causality of AD are rooted in the past in the sense that: (1) amyloid and tau biomarkers have been highly associated with disease, (2) genes encoding amyloid and tau pathways can cause AD, and (3) treatments targeting these pathways have some effects on symptoms. However, modern epistemic theories of causality recognize that these biomarkers may only be tangentially related, may not be required for causation, and that multiple other factors are also causal. Synaptic and neuronal loss results from multiple insults, including those resulting from upstream inflammation, lack of cardiovascular support, and metabolic dysfunction. Understanding of the pathologies that lead to Alzheimer’s dementia is improving but incomplete. However, it is increasingly clear that the number of involved molecular domains and the severity of their involvement increases over time; treatment should begin as early as possible in AD. In particular, those at the highest risk—for example, based on genetic screening—should initiate preventive treatment as early as possible. In all cases, treatment should start before the onset of dementia. Our recommendations emphasize treatment of AD at its earliest stages, ideally presymptomatic. Individuals should be screened for risk, and those at highest risk should prioritize early intervention. However, if the early treatment window is missed, later treatment should not be denied. AD interventions are likely to have at least some effect at all stages of disease, but we recognize AD as a continuum with on-going recruitment of additional disease perturbed biological networks, so interventions that are effective in early stages of AD are likely to be less effective in later stages.

As the disease progresses, the specifics of certain interventions (like exercise) might have to be modified (e.g., for exercise: type, duration, frequency, setting, intensity, amount of caregiver assistance, required cognitive load), but the underlying value of targeting the molecular pathways influenced by these interventions is likely to remain useful.

### AD has multiple causes

A ***cause*** of a disease is most strictly defined as *a specific factor without which the disease would not occu*r (e.g., Rothman, 1976). This definition works well for diseases with simple linear chains of causation (**Figure 1A**) and can even be adapted if there are multiple distinct possible causes converging on a single path (**Figure 1B**). A more pragmatic definition of ***cause*** is *<u>any factor that brings about change for better or worse in a health condition</u>* (Smith & Susser, 2002; Kaufman & Poole, 2000). For purposes of conciseness and clinical relevance, we embrace this pragmatic definition, which encompasses both deterministic molecular pathways and modifiable risk factors (**Figure 1C**). However—where possible—we distinguish between molecular causes and risk factors because (1) advances in precision & personalized medicine benefit from a mechanistic understanding of the relationships between risk factors and molecular pathways, and (2) modifiable risk factors vary in effect size and clinical importance whereas molecular pathways are deterministic. A reason to favor the more general definition of cause is that more than one deterministic molecular pathway may lead to AD. For example, it might be that either inflammation alone or amyloid accumulation or metabolic insufficiency alone could cause AD. Or—in many individuals—a combination of multiple molecular causes could drive AD onset and progression. Thus, it is important to avoid a paradigm that invites envisioning the cause of AD as ultimately confined to—or channeling into or through—a single path. With this conceptualization, it becomes easy to surmise that subgroups of AD arising from different molecular causes will likely require different treatments targeting different domains of intervention.

**Figure 1.**
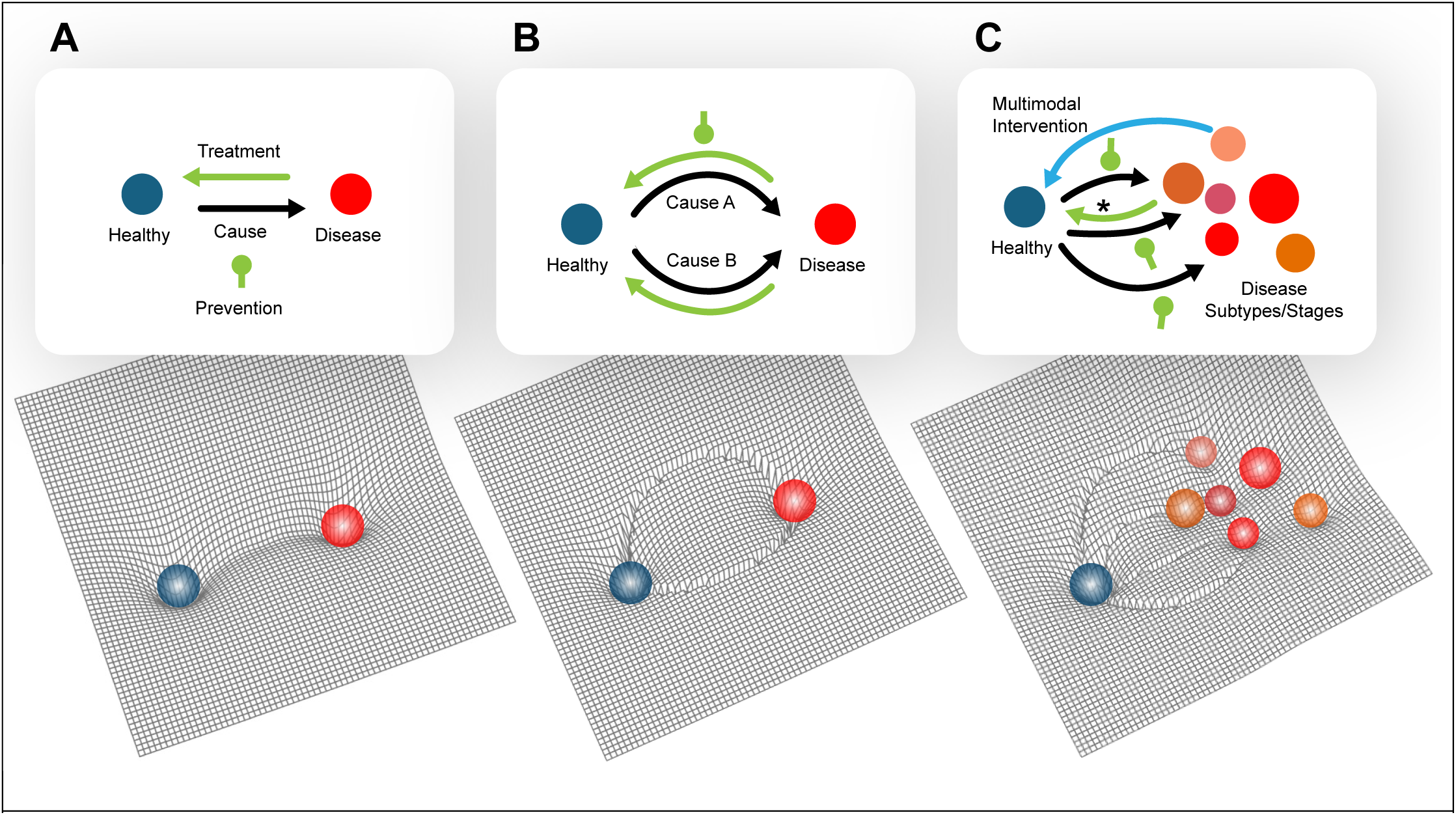
Alzheimer’s disease is a complex disease requiring complex treatments. Conceptualization of an “energy landscape” for AD. The *x* & *y* coordinates are shorthand for the millions of variables that represent the current status of the human biosystem. The slope of a path in the landscape can be considered the probability that an individual will move in that direction in a given unit of time. (A) *Classic concept of disease*: The system moves (black arrow) from a healthy state through a single obligatory causal state (e.g., head trauma) to a state of disease defined by clinical symptoms (e.g., chronic traumatic encephalopathy / CTE). Treatment consists of blocking (green club) or reversing the transition of the system along a linear path (green arrow). (B) *Extended classic concept of disease*: The system moves from a healthy state through one of several possible causal states (e.g., Paraquat toxicity or GBA genetics) to a state of disease (e.g., Parkinsonism). Treatment consists of blocking or reversing the transition of the system along one of the causal trajectories, as diagnosed in a particular individual. (C) *Complex disease*: The system moves from a healthy state through potentially multiple possible causal states to one or more similar but not identical subtypes, which in ensemble form a diagnostic category (e.g., Alzheimer’s disease), possibly transitioning or progressing between different states. Treatment consists of preventing the system moving towards unhealthy states, and/or moving the system towards the healthy attractor, possibly along different paths (blue arrow) than those used to reach the disease states (*). Multidomain therapies can work simultaneously to move the system along these favorable multidimensional trajectories.

An alternative lexicographical viewpoint would be to define any clinical entity that cannot be understood as the result of single unbranched chain of causality not as a unique *disease*, but rather a *syndrome*. Such a syndrome, after sufficient future research, would eventually be reclassified as several separate diseases (Peterson & Keeley 2015). This may ultimately be true for AD, but we remain skeptical that resolving what we believe to be a web of factors that synergistically interact into separate definable diseases will ever be clinically practical.

Furthermore, in practice, the momentum of historical usage of the phrase “Alzheimer’s disease” is so strong that it may never be renamed (Whitehouse & George, 2015). Therefore, we are comfortable in our description of AD as a *disease with multiple causes*. Other complex diseases (or syndromes?) such as Parkinson’s disease (PD) face similar classification issues; Espay and Lang (2018), for example, recognize that omics and other objective tests may assist in classifying subtypes of PD. They use a metaphor of duck attributes: every duck is unique but together they form a recognizable species (Figure 1B from Espay and Lang).

The multiple causes of AD have been amply reviewed (e.g., Gong et al., 2022; Huang & Mucke, 2012; Zhang et al, 2018; Wainberg et al., 2021; Ferrer, 2022; Masurkar et al., 2024). There remain differences of emphasis by different stakeholders on whether and to what extent these causes merge onto a final common pathway (e.g., Delport & Hewer, 2022). Our approach to AD translational research is valuable for all proposed models.

### AD can be categorized into *subtypes*

Every individual with AD has their own set of causes of AD. In clinical practice, it is convenient to cluster groups of patients with a similar set of causes as a ‘subtype’ of AD (e.g., Kikuchi et al., 2022); these individuals would occupy the same attractor state (or “well”) modeled in **Figure 1C**. These states are more precisely defined by molecular sequelae (e.g., biomarkers) than causes, but the relationships between causes and sequelae (if measured with omics) should be approximately a bijection, so these perspectives are interchangeable. Systems approaches, in addition to *defining* AD subtypes, may be particularly useful in *identifying transitions*, or ‘tipping points’, into or between these subtypes (e.g., Aihara et al., 2022; Liu et al., 2020; Chen et al., 2017). This systems view has implications for translational research and the design and refinement of AD therapies—whether lifestyle or pharmaceutical (Uleman et al., 2024). A therapeutic target (“drug target”) should not be a specific enzyme or molecule—or even a pathway—but rather should be *a network or a set of interacting networks*. This paradigmatic shift may justify greater conceptual emphasis on broad lifestyle interventions such as diet and exercise that may be highly effective but are better described as targeting networks than molecules. Each subtype may be modeled as an attractor state, and stages of disease may be modeled as a series of attractor states linked by transitional states (**Figure 1)**. Taherian Fard and Ragan (2017) illustrate a method for quantifying the “energy” of such a landscape based on correlations of omics measurements and provide an example for PD. Statistical learning from omics data informs unbiased clustering of AD cases into subtypes (Neff et al., 2021; Higginbotham et al., 2023; Tijms et al., 2024); such subtypes likely correspond to attractor states. Tijms et al. ascribe potential causes to five subtypes based on enrichment of analyte function metadata: hyperplasticity, innate immune activation, RNA dysregulation, choroid plexus dysfunction, and blood–brain barrier dysfunction; these likely correspond to a different mix of causes for cases in each cluster. Individuals with different risk factors are likely to be enriched in different molecular clusters corresponding to the causes of their AD. For example, specific genotypes—such as APOE4 homozygosity—may drive individuals towards particular attractor states (Arnold et al., 2020; Fortea et al., 2024).

### AD can be categorized into *stages*

Somewhat orthogonally to the concept of clustering cases by molecular <u>causes</u>, one may cluster cases by the <u>stage</u> of disease. A given individual with the same set of causes will likely transition through multiple stages over the course of their disease. If these stages are sufficiently distinct, they may be defined by attractor states (**Figure 1C**); if not, progression occurs smoothly along subtle contours of the modeled topography, and “staging” represents arbitrary but convenient thresholds. Historically—and to-date the most practical—methods of AD staging are based on cognitive assessment. A patient with AD but no symptoms is *presymptomatic*. A patient with complaints (perhaps brain fog, amnestic complaints, or executive function complaints) but who still scores normally on neuropsychological tests has *early AD* (and perhaps labeled with the deprecated term “subjective cognitive impairment”, or SCI). A patient with abnormal neuropsychological testing with preserved activities of daily living (ADLs) has a *mid-stage AD* (and perhaps labeled with the deprecated term “mild cognitive impairment”, or MCI—even though there may be severe molecular dysfunction at such a stage). A patient with Alzheimer’s dementia has *late-stage AD*. Use of this historical approach to staging impairs consistency (not only between providers but also for the same provider across decades), as thresholds separating the stages are subjective and/or determined the type and sensitivity of neuropsychological test(s). FAST staging (Reisberg et al., 2010) is one such approach: stage 1 reflects presymptomatic disease, stage 2 reflects SCI, stages 3 reflects MCI, and stages 4-7 reflect increasingly severe dementia. The CDR (Lanctôt et al., 2024) provides a similar framework but lumps early stages (presymptomatic, SCI, and MCI) together in stage 0, so is less useful. The CDR’s resolution can be improved by utilizing “sum-of-boxes” to get the CDR-SB (Tzeng et al., 2022). Improved molecular, imaging, and neurocognitive tests should soon enable more precise and consistent staging, particularly in presymptomatic individuals. One area of future research is to clarify whether stages represent distinct attractor states, or whether they are arbitrary designations based on thresholds of a continuum.

### Each of the molecular causes of AD may be driven by multiple modifiable risk factors

Proposed molecular drivers of AD include accumulation of amyloid β protein (Aβ) in neuritic plaques—or, more generally, disrupted proteostasis, disruption of the astrocyte-neuron lactate shuttle, and many other mechanisms (e.g., Paterson et al., 2024; Zhang et al. 2023). Cardiovascular and metabolic dysfunction are two examples of modifiable causes of AD at a physiological systems level. These systems-level causes may each influence one or more specific molecular causes. Together, these encompass multiscale networks that influence molecular dysfunction of neurons and their synapses, and drive AD. Diet and exercise ***both*** influence ***multiple*** molecular system causes. Lifestyle and pharmaceutical interventions can be roughly categorized into domains that are generally more descriptive of the intervention (e.g., diet) than the physiologic and molecular systems they target. An oversimplification would be that each intervention domain corresponds 1:1 to a physiologic or molecular domain, but that is unlikely to be the case for most lifestyle interventions.

### Muti-domain or -modal?

We use the concept of “domain” to refer to categories of molecular subsystems, such as inflammation, proteostasis, cardiovascular, or metabolism. Therefore, a multidomain intervention is one that targets multiple subsystems (aka, “pathways”). We use the concept of “mode” to refer to a particular intervention, such as a particular pharmaceutical or diet. Most multimodal interventions are also multidomain interventions, but not necessarily, as multiple modes could conceivably all target the same domain. A mode of intervention (where “mode” is a descriptive classification of the type of intervention—such as “dietary”) could impact multiple molecular system domains. For example, exercise targets multiple domains. For reasons of conciseness, even though trials using only exercise as an intervention are therefore formally “multidomain”, we do not thoroughly review them here (e.g., they are not included in **Table 2**). “Multimodal” (or sometimes “multicomponent”) is used to indicate multiple types of intervention, even if they are targeted to the same domain (Abramowitz & Weber, 2024). ‘Multimodal’ and ‘multidomain’ are often confused, and in the literature are generally conflatable, along with ‘multidimensional’, ‘multisystem’, and ‘multi-domain’. None of these terms is synonymous with “lifestyle intervention”; a multidomain or multimodal intervention could consist entirely of pharmaceuticals, entirely of lifestyle interventions, or a mix of both. Current frameworks for quantifying the impact of particular modes of intervention (e.g., moderate aerobic exercise or coaching support for a Mediterranean diet) on particular molecular domains are insufficient (Soldevila-Domenech et al., 2025). Indeed, a precise delineation of each domain is also currently insufficient. Using omics together with other objective measures (e.g., imaging or neurocognitive markers) to better define domains and to measure changes in these domains in response to modes of intervention is a current research priority. We also recognize that some subfields use “mode” terminology to differentiate broader categories of intervention; notably in the context of exercise science, “mode” may refer to a particular type of exercise performed at a particular intensity. Our use is consistent with that terminology, but we would most likely not consider an intervention consisting solely of multiple modes of exercise to be multi<u>domain</u>.

## METHODS

### Scoping Review Objectives

Although the claim may seem obvious that there is a need for multidomain treatment of patients with AD, most clinical guidelines currently only give lip service or short shrift to multidomain interventions. A compilation of studies clarifies gaps in research and provides a resource to the community. Much discussion has been made of multidomain interventions, but often in the context of partial reviews of the literature (Lin et al., 2016). Soldevila-Domenech et al. (2025) provide one compilation, but their compilation skips some studies we include, and focuses on trial design, not on trial results. Also, their compilation focuses on the concept of “dose” of a multidomain therapy, so includes studies not related to AD. **Scoping Review Methods**. A Preferred Reporting Items for Systematic reviews and Meta-Analyses extension for Scoping Reviews (PRISMA-ScR) checklist is included as **Supplemental Material** (Tricco et al., 2018).

#### Identification of relevant studies

PubMed was queried for relevant studies with criteria crafted to include three conceptual components: (1) Alzheimer’s disease (any stage along the spectrum), (2) clinical trial, and (3) non-pharmacological multidomain intervention. The exact Boolean and resulting flow chart are provided in

### Supplemental Appendix 2

#### Inclusion/exclusion criteria

Out of all potentially relevant publications, those meeting our study selection criteria were retained. Selection criteria were:

1. Publication available in English in a peer-reviewed journal with full text availability.
2. Inclusion criteria designed such that a majority of participants would be expected to have AD (any stage, including presymptomatic) at the time of enrollment.
3. Trial enrollment is not restricted to DIAD.
4. Prospective RCT.
5. At least one primary outcome is a valid quantitative measure of function or cognition.
6. Intervention included at least two non-pharmacological modalities spanning at least two domains of intervention.
7. At least two arms, one of which is a control.
8. Trial is completed, and results for primary outcome(s) are published.

Studies not quite meeting inclusion criteria, and the reasons for exclusion, are listed in **Supplemental Appendix 2**. In order to avoid missing high-quality systematic reviews and RCTs, the PubMed FFT (free full text) filter was <u>not</u> used (Krieger et al., 2008). The period of the literature search was all dates prior to June 13, 2025.

#### Unpublished (ongoing) studies

Separately from the scoping review analysis, unpublished ongoing studies were non-systematically identified by searching Google Scholar and ClinicalTrials.gov using the same three conceptual components of the scoping review. These studies are included either in **Table 1** labeled with an asterisk (if they meet scoping criteria 1,2,3,5, and 6) or **Supplemental Appendix 2** for context.

**Table 1.**
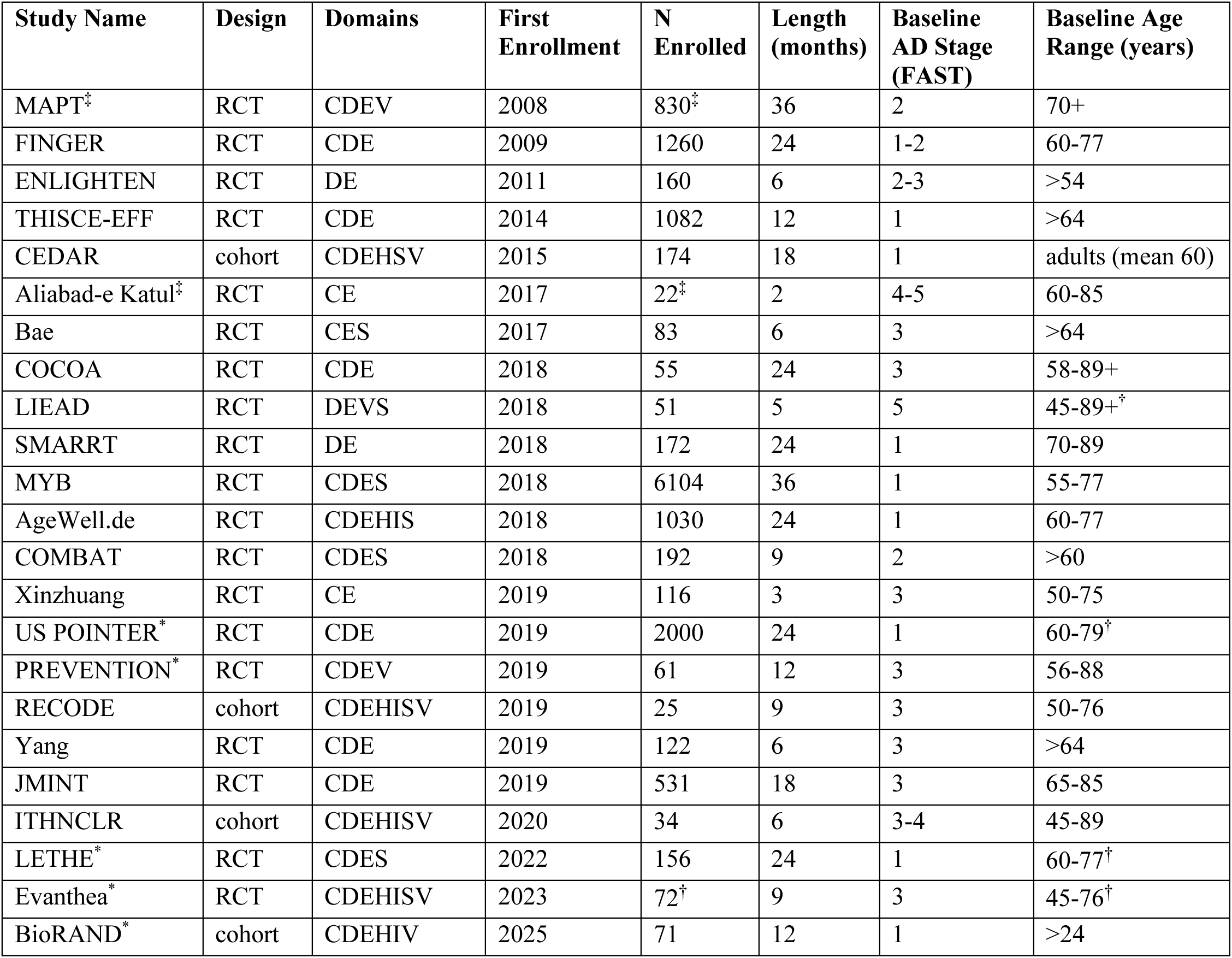
Data Extraction Sheet for studies informing multidomain interventions. Staging is reported as best estimate from reported trial inclusion criteria (**Supplemental Appendix 1**). Trials without established names/acronyms are referred to by their geographic location. References: AgeWell.de (Zülke et al., 2024); Aliabad-e Katul (Damirchi et al., 2018); Bae (Bae et al., 2019); BioRAND (Niotis et al., 2025); CEDAR (Isaacson et al., 2019; Saif et al., 2022); COCOA (Roach, Edens, et al., 2022; Roach et al., 2023); COMBAT (Liu et al., 2023); ENLIGHTEN (Blumenthal et al., 2019); Evanthea (ClinicalTrials.gov ID NCT05894954); FINGER (Ngandu et al., 2022); ITHNCLR (Sandison et al., 2023); JMINT (Sakurai et al., 2024); LETHE (Rosenberg et al., 2024); Lifestyle Intervention for Early Alzheimer’s Disease (LIEAD) (Ornish-D et al., 2024); MAPT (Andrieu et al., 2017); MYB (Brodaty et al., 2025); PREVENTION (McEwen et al., 2021); RECODE (Toups et al., 2022; ClinicalTrials.gov ID NCT03883633); SMARRT (Yaffe et al., 2023); THISCE-EFF (Chen et al., 2020); US POINTER (Baker et al., 2024); Xinzhuang (Fan et al., 2024); Yang (Yang et al., 2022). *In progress / final results not yet published. ^†^Planned. NR=not reported. Domains (for details see individual trial publications as these are approximate): C = cognitive training; D = diet/metabolism; E = exercise; H = endocrine; I = inflammatory/infectious; V = vitamins/supplements; T = toxin reduction; S = social/psych. ^‡^more than two arms; some arms included multidomain interventions.

| Study Name | Design | Domains | First Enrollment | N Enrolled | Length (months) | Baseline AD Stage (FAST) | Baseline Age Range (years) |
| --- | --- | --- | --- | --- | --- | --- | --- |
| MAPT<sup>‡</sup> | RCT | CDEV | 2008 | 830<sup>‡</sup> | 36 | 2 | 70+ |
| FINGER | RCT | CDE | 2009 | 1260 | 24 | 1-2 | 60-77 |
| ENLIGHTEN | RCT | DE | 2011 | 160 | 6 | 2-3 | >54 |
| THISCE-EFF | RCT | CDE | 2014 | 1082 | 12 | 1 | >64 |
| CEDAR | cohort | CDEHSV | 2015 | 174 | 18 | 1 | adults (mean 60) |
| Aliabad-e Katul<sup>‡</sup> | RCT | CE | 2017 | 22<sup>‡</sup> | 2 | 4-5 | 60-85 |
| Bae | RCT | CES | 2017 | 83 | 6 | 3 | >64 |
| COCOA | RCT | CDE | 2018 | 55 | 24 | 3 | 58-89+ |
| LIEAD | RCT | DEVS | 2018 | 51 | 5 | 5 | 45-89+<sup>†</sup> |
| SMARRT | RCT | DE | 2018 | 172 | 24 | 1 | 70-89 |
| MYB | RCT | CDES | 2018 | 6104 | 36 | 1 | 55-77 |
| AgeWell.de | RCT | CDEHIS | 2018 | 1030 | 24 | 1 | 60-77 |
| COMBAT | RCT | CDES | 2018 | 192 | 9 | 2 | >60 |
| Xinzhuang | RCT | CE | 2019 | 116 | 3 | 3 | 50-75 |
| US POINTER<sup>*</sup> | RCT | CDE | 2019 | 2000 | 24 | 1 | 60-79<sup>†</sup> |
| PREVENTION<sup>*</sup> | RCT | CDEV | 2019 | 61 | 12 | 3 | 56-88 |
| RECODE | cohort | CDEHISV | 2019 | 25 | 9 | 3 | 50-76 |
| Yang | RCT | CDE | 2019 | 122 | 6 | 3 | >64 |
| JMINT | RCT | CDE | 2019 | 531 | 18 | 3 | 65-85 |
| ITHNCLR | cohort | CDEHISV | 2020 | 34 | 6 | 3-4 | 45-89 |
| LETHE<sup>*</sup> | RCT | CDES | 2022 | 156 | 24 | 1 | 60-77<sup>†</sup> |
| Evanthea<sup>*</sup> | RCT | CDEHISV | 2023 | 72<sup>†</sup> | 9 | 3 | 45-76<sup>†</sup> |
| BioRAND<sup>*</sup> | cohort | CDEHIV | 2025 | 71 | 12 | 1 | >24 |

#### Data extraction

For each study, we extracted the study name, design (RCT or cohort), domains of intervention, first enrollment date (year), number enrolled (intent to treat), length (months), baseline AD Stage (FAST), baseline age range (years), primary outcome measure(s), effect size (relative difference and/or Cohen’s d), and primary outcome measure significance (p-value). Data was interpreted only from published papers and ClinicalTrials.gov records; no attempt was made to obtain or confirm data from investigators. The data extraction sheets are reported in **Table 1** and **Table 2**.

**Table 2.** Results of multidomain intervention RCTs (trials included in scoping review). These studies are sufficiently diverse in design (e.g., with respect to population, outcome measure, nature of control arm, length, and intervention) as to discourage meta-analysis. No trials share any primary outcome measure (as composite scores are bespoke to each trial). Relative difference is between arms at trial endpoint. P-values are raw reported for multidomain arm vs control arm. Cohen’s d values are best estimates from reported trial data. MOCA_adj_ is a Taiwanese version of MoCA including a one-point addition for education <12 years. ∞ = improvement (or no change) in intervention compared to decline in control; *both arms <u>improved</u> with respect to baseline. ^†^Only one of six NCGG-FAT cognitive measures was significantly different. ^‡^MAPT arm 1 included omega-3 supplementation with other domains of interventions; arm 3 did not.

| Study Name | Primary outcome measure | Trial Duration (mos) | Relative difference (%) | Effect size (Cohen's d) | Significance (p-value) |
| --- | --- | --- | --- | --- | --- |
| MAPT arm 1 | Composite Z-score | 36 | ∞ | 0.09 | 0.047 |
| MAPT arm 3 | Composite Z-score | 36 | ∞ | 0.08 | 0.090 |
| FINGER | Composite Z-score | 24 | 25 | 0.13 | 0.03 |
| ENLIGHTEN | Ranked composite | 6 | ~21 | 0.40 | 0.012 |
| THISCE-EFF | MOCA<sub>adj</sub> | 12 | ∞ | 0.2 | 0.094 |
| Bae | NCGG-FAT battery | 6 | ∞ | 1.5 | 0.003<sup>†</sup> |
| COCOA | MPI score | 24 | 60 | 3.2 | 0.0007 |
| COCOA | FAST | 24 | 38 | 1.8 | 0.03 |
| LIEAD | ADAS-cog | 5 | ∞ | 1.2 | 0.053 |
| LIEAD | CDR-SB | 5 | 15 | 0.68 | 0.032 |
| SMARRT | Composite Z-score | 24 | 74 | 0.14 | 0.02 |
| MYB | Composite Z-score | 36 | 280* | 0.18 | 0.001 |
| AgeWell.de | Composite Z-score | 24 | 19* | 0.01 | 0.87 |
| COMBAT | Composite Z-score | 9 | 948* | 0.40 | 0.026 |
| Xinzhuang | MMSE | 3 | ∞ | 0.28 | 0.003 |
| Yang | MoCA | 6 | ∞ | 1.6 | 0.022 |
| JMINT | Composite Z-score | 18 | ∞ | 0.09 | 0.23 |

### Dominantly inherited AD (DIAD)

About 1% of cases of AD are DIAD. Strong evidence using direct logical approaches supports each genetic cause of DIAD as a distinct disease. This evidence includes highly penetrant dominant inheritance with rare cases of single epistatic genes that block a single disease-causing pathway (**Figure 1A**). These DIAD diseases are solely due to variation in a single gene and are therefore not complex diseases. As such, our arguments for synergistic multidomain therapy are weaker for DIAD patients. However, similarities between DIAD and other cases of AD—both across symptoms and molecular pathophysiology— particularly in symptomatic stages, may support a role for multidomain therapy in DIAD.

## RESULTS

### Patients with AD need multiple interventions

Norton et al., (2014) identified seven modifiable risk factors (which are also “causes” by the definition we are using in the present context) accounting for 30% of AD cases: low education, midlife hypertension, midlife obesity, diabetes, physical inactivity, smoking, and depression. In 2020, the Lancet Commission expanded these to twelve modifiable risk factors accounting for 40% of AD cases, adding: hearing impairment, low social contact, excessive alcohol consumption, traumatic brain injury, and air pollution (Livingston et al., 2020). Additional modifiable causes continue to be identified, such as untreated vision loss and high mid-life cholesterol (Livingston et al., 2024). The emergence of multidomain interventions for AD derived in large part from this evidence of modifiable risk factors.

Since AD has multiple etiologies, it makes sense that treatment —whether preventive, symptomatic, or curative—should target multiple domains. The best treatment is likely to be synergistic (Uleman et al., 2023). Molecular systems are highly intertwined and synergistic; improving function in one domain may improve function in other domains. Efforts to treat AD via a single domain—consider donepezil—have occasionally transiently and marginally improved function (e.g., Birks & Harvey, 2018), but none have had the impact of multidomain interventions (**Table 2**). Most studies of multidomain interventions for AD show a significant and substantial effect on cognition. Of the 14 completed trials, only 3 (THISCE, JMINT, and AgeWell.de) do not show significant beneficial results, although three additional trials (Bae, LIEAD, & MAPT) with multiple arms or primary outcome variables did not show significance in all of these.

### The “placebo” effect

AgeWell.de, MYB, and COMBAT showed improvements in both the intervention and the control arm **(Table 2)** suggesting that the controls received substantial lifestyle benefit during the trial period and may have succeeded in ameliorating AD on their own. Such an effect in the context of a pharmaceutical trial might be considered a placebo effect, but it is likely that individuals who enroll in lifestyle intervention trials will change their lifestyle regardless of what arm they are assigned to or the intention of the trialists to partition intervention. Even simple interactions with trial staff may provide a modicum of “social” intervention. Control participants may be motivated to exercise on their own or pursue dietary health.

Enrollment in a trial may reward altruism with improvements in mood. Such effects are likely to dampen significances and effect sizes in real-world multidomain trials compared to a gedanken trial with a theoretically perfect control arm, and likely explain improvements seen in some control arms. We conclude that many multidomain trials had less power than may have been expected. Addition of multi-omic measurements (Diener et al., 2025) to future trials will be critical to provide precise quantitative measurements of actual dose and exposure and effect on molecular systems received by all individuals in a trial (regardless of arm) to enable better interpretation of trial data and improved statistics.

### Controlled Trials and Cohort Studies that Inform Multidomain Interventions

Although some studies have looked at single interventions based on these risk factors (e.g., physical activity, cognitive training, or aggressive hypertension control), the results have not been compelling. The lackluster success of unimodal interventions was the impetus for studies of multimodal, multidomain interventions (**Table 1**), including the now well-known Finnish Geriatric Intervention Study to Prevent Cognitive Impairment and Disability (FINGER) trial in older adults at risk for cognitive decline (Ngandu et al., 2022). We summarize multidomain RCTs for AD in a data extraction sheet in **Table 1**. Extraction of metadata for these trials is described in **Supplemental Appendix 1**. Related studies not meeting criteria for inclusion in **Table 1** are summarized in **Supplemental Appendix 2**. FINGER showed that a 2-year multidomain intervention focused on diet, exercise, cognitive training, and management of cardiometabolic risk factors significantly improved neuropsychological functioning compared to a control group that received general education. The benefits of the intervention were seen irrespective of baseline factors including sex, age, education, socioeconomic status or cardiometabolic risk factors (Rosenberg et al., 2018), although ethnic diversity was limited in this cohort. The success of FINGER has led to a global effort to expand on the findings, World-Wide FINGERS (Rosenberg et al., 2020; Kivipelto et al., 2020). A meta-analysis of 28 studies of multidomain interventions compared to single interventions in >2700 patients with mild cognitive impairment (MCI) found that multidomain interventions resulted in significantly greater improvements in global cognition, executive function, memory, verbal fluency and other cognitive measures (Salzman et al., 2022).

Although all of these trials have generally consistent results, most with substantial improvement for those receiving multidomain interventions, there are numerous differences between the trials and their results. Quantitative meta-analysis would be ill advised. Some randomized controlled trials (RCTs) focus recruitment on individuals with presymptomatic AD, typically by identifying “at risk” individuals or individuals with cognitive test scores slightly below average, but still “normal”. Other RCTs focus recruitment on early AD, with individuals with objectively low cognitive test scores, but typically no loss of daily living function. Many of these early AD individuals would under previous nomenclature and classification systems have been classified as having MCI (Petersen et al., 2014; Bermejo-Pareja & Del Ser, 2024). No studies have focused on individuals with late AD (Alzheimer’s dementia), but some late AD individuals have been included in some of the RCTs (notably LIEAD, **Table 1**), as inclusion criteria are typically based on test-score thresholds and not presence/absence of dementia. Although these studies have limitations, there is no obvious trend of reduction of benefit in presymptomatic compared to early AD, and since there is no absolute reason why demented individuals would be completely unable to receive multidomain interventions—they can exercise, they can change their diet, they can take medicines—we would expect by extrapolation that at least some benefits would accrue at all stages of AD. However—in our subjective experience as clinicians—it becomes nearly impossible to introduce meaningful intervention in late stages of dementia, emphasizing the desirability of early intervention. One criticism of multidomain treatments is the lack of data from patients with Alzheimer’s dementia (as opposed to earlier stages of AD). Indeed, any potential value of multidomain treatments—or any treatment—in patients with more advanced dementia needs to be tested in clinical trials—but not at the expense of denying this care currently to patients with presymptomatic or less advanced disease. The effectiveness of multidomain treatments for prevention and treatment of early-stage disease seems clear.

In **Table 1**, trial size (N) and length of trial are positively correlated, and both are negatively correlated with baseline FAST (**Supplemental Figure 1 & 2**). These relationships support the perhaps commonsense hypothesis that logistics for large trials are easier for early-stage than for late-stage AD, particularly with regard to cost-per-participant, ease of recruitment, and ease of long-term follow-up. No relationships are observed between outcome significance or strength and any of trial size, trial duration, or baseline FAST. One would be reluctant to draw conclusions from these observations of lack of relationships, as the data from the observed trials only thinly samples all possible trial implementations, but these observations may hint that all of these factors together contribute to trial power, and that stakeholders designing or evaluating trials should consider all of these factors together in power analyses rather than focus on N alone. It remains unproven that (1) large AD trials are more powered than small AD trials, (2) late-stage trials are more powered than early stage, or that (3) longer trials have more power. Assumptions that these three statements are true should therefore be used with caution during trial design.

In light of these considerations, trial designers must navigate between the Scylla of real-world clinical desirability of intervention at a pre-symptomatic stage and the Charybdis of funding and logistical need for a short trial with classical statistical power to see a strong, significant result for a pre-declared endpoint—which in the last century required shorter trials with highly impaired individuals more likely to significantly decline in a control arm. Modern Odysseus-like trial designers should navigate this strait by using omics-based trials to observe early molecular changes in presymptomatic AD, thereby focusing on individuals most likely to benefit, with the most effect size, in a short time, using massive data, but avoiding the expenses, logistics, and risks— and perhaps impossibility—of recruiting many AD patients for years-long trials. Trials may leverage large, shared pools of potential participants such as ALZ-NET (2025), and/or could explore novel methods of fully remote recruitment. Furthermore, participant baseline attributes, degree of adherence/compliance with the intervention, treatment received by controls, and other methodologic factors may account for some differences between multidomain intervention results. Therefore, future research in multidomain interventions should be able to comprehensively measure the system of each participant (including those in controls) both at baseline and throughout the period of study. Measures of adherence/compliance should include both non-molecular (Soldevila-Domenech et al., 2025) and molecular measures (Roach, Hodes, et al., 2022). These trials should be modernized with dense data designs, as we argue below.

### Excuses not to act

Futility has been suggested as a roadblock to action. Reasons for inaction arise in many forms, including: (1) “People with dementia cannot exercise”; (2) “The level of proof is not sufficient”; (3) “Costs are not reimbursable”; (4) “I just want to take a pill”; (5) “I don’t know exactly what the best diet is, or the best course of exercise, so I won’t even try a potentially suboptimal intervention.” (6) “I want to start with just one thing, then add other interventions if it does not work”. These crutches of inaction illustrate many barriers, none of which are insurmountable, by individual patients, caregivers, clinicians, and/or society.

Education will be key to overcoming these barriers. Education is needed at all levels: policymakers, providers, caregivers, and patients. Coaching, whether by AI or in person, is likely to be an effective form of personal education. The very nature of multidomain interventions makes them exceptionally flexible. If an intervention (e.g., a gym membership) is too expensive, find another (e.g., a walk around the block) that is less expensive. If good intervention choices cannot be easily identified, leverage human or AI coaching. Perfection is the enemy of the good; any healthy diet is likely to be better than none (e.g., Pontzer, 2021). There is not time to iterate over each possible single domain. Iterating interventions is good where possible, such as in hypertension control. But for AD, waiting months or years to see the effects of each intervention in isolation will take too long – the disease will progress, healthspan will be significantly shortened. From a clinician’s point of view, and therefore a patient’s, multidomain intervention must start at diagnosis (or possibly before, based on risk factors), and be based on best available evidence, even if the perfect intervention may remain uncertain.

### Need to Personalize Multidomain Interventions

There are multiple causes of AD. Multiple interventions may ameliorate AD. Different individuals with AD will have different mixes of causes. Therefore, personalized treatment will be needed (Niotis et al., 2025). These concepts are illustrated in **Figure 1**. One of the challenges in implementing multidomain interventions, and in analyzing and interpreting their outcomes, is the variability in intervention design, including intervention length, treatment “dose” (i.e., number and frequency of sessions), and types of interventions included. Even within a trial, exact details of the multidomain intervention will differ between individuals. For example, if a goal is to reduce blood pressure across all participants receiving a multidomain intervention, the exact drugs and lifestyle changes to reduce pressure should vary between individuals. Even for fixed-dose pharmaceuticals, compliance, bioavailability, and half-life may vary between individuals. To address the need for scientific consistency in the presence of real-world variability, Solomon et al. (2021) have proposed a model for “Brain Health Services” which allows for a degree of standardization while still personalizing the intervention to an individual’s specific clinical needs and motivation to change lifestyle behaviors. Nutrition interventions for AD prevention or treatment may be considered “personalized medicine” because many nutrients target specific pathways that have been shown to be altered in the disease. For example, a Mediterranean-type diet provides higher intakes of nutrients such as omega-3 fatty acids (from cold water fish), B-vitamins which lower homocysteine levels (from green vegetables and whole grains), and altered patterns of dietary fat types which can influence blood levels of lipids and phospholipids related to AD risk. This has been the basis of several personalized nutrition approaches to reducing cognitive decline in individuals at increased genetic risk for AD or with early-stage cognitive decline (Norwitz et al., 2021; Amini et al., 2020).

If our underlying and overarching hypotheses are true — that AD has multiple causes and is best treated with multiple interventions — then conclusions and clinical recommendations resulting from such trials are likely to be applicable to all individuals regardless of where they are on the AD spectrum. We may be able to extrapolate conclusions from such trials to many real-world patients. For example, perhaps with help of caregivers, even individuals with advanced dementia are capable of modifying their diet, most are capable of exercising, and pharmaceutics can be optimized for all. Multidomain recommendations can be adapted to the specific circumstances of each individual. It is also possible to classify each mode of intervention as either “general” (applicable to many conditions of health or disease, such as exercise) or “AD-specific” (applicable only to AD or very similar disease, such as donanemab). Such classification may help in didactics, epistemics, developing guidelines, and allocation of responsibilities between providers.

## CONCLUSIONS

### Personalization requires measurement

Advances in science are often dependent on advances in instrumentation, or the ability to measure things. To advance understanding and treatment of a systems disease such as AD, advances in measuring systems are needed. Such advances have happened in recent years, and continue to accelerate, notably by bringing down the cost and increasing the scope of multiomics. Advances in imaging and molecular biomarkers are also transforming AD care and research (e.g., Soldozy et al., 2019). Key needs for future advances will include optimizing signal-to-noise for measurements of system function and tracking the “dose” of each domain intervention. Most if not all lifestyle interventions are not precisely defined or quantified. There are infinite personal variations to food choice, timing, and amount even for concepts with specific names such as “Mediterranean diet” (Tessier et al., 2025). This vagueness extends to cognitive training and exercise domains. Some domains, such as sleep, are reasonably quantifiable; we need to bring similar quantification to all domains.

### Research requires measurement

Iterative refinement is key not only to clinical care of individuals but also to design clinical studies of the complex interventions that must be central to research-based guidelines for treating complex diseases such as AD. Iterative refinement of these complex interventions—such as diet and exercise— cannot be made until their effects can be objectively measured at the physiologic and molecular levels. This will require understanding these systems and their networks sufficiently enough so that quantifiable objective biomarkers—likely blood based—can be used to measure the impact of each intervention in each person on each system. Until these measurements are in place, studies and clinical trials will remain incomparable (**Table 2**). Meta-analyses will remain out of reach; validation studies will not yield insights into future trial designs.

The ensemble of these omic, imaging, psychometric, and other data can be termed “systems quantification”. In addition to the value to research, systems quantification will also guide therapy, as summarized above (Uleman et al., 2023). A key element of systems quantification—one that is easily in reach of modern studies—will be to quantify compliance of study participants both with global and with specific interventions. Understanding compliance will enable interactive improvements in personalized recommendations. Soldevila-Domenech et al. (2025) have suggested merging multiple domains into a single-dimensional compliance metric. We believe that a single metric of compliance could be valuable as a global indicator of the logistical performance of the trial infrastructure and implementation. However, because interventions and responses are multidomain and individual, a vector of compliance values will be necessary in order to advance understanding. Conversion of this vector of values to a global value should vary from individual to individual based on the particular nature of that individual’s needs to maintain and/or return to homeostasis. Furthermore, we anticipate that the most useful metrics will be objective and quantifiable and tied closely to molecular mechanism (such as molecular biomarkers of physiologic processes). These are likely to be more tightly correlated to causal influence on outcome measures than factors such as number of visits to a provider or amount of time engaged with cognitive training, due to the large number of factors (e.g., attention, alertness, exact nature of the intervention, personal traits, and many others) that modify or influence these measures on health outcomes. For example, diet questionnaires are nearly useless for understanding the impact of a particular diet change on a patient’s metabolism, but quantifiable molecular markers may highlight key successes and opportunities for improvements. Improvements in multidomain interventions will remain glacial until these domains are standardized by quantifiable biomarkers. Quantifiable biomarkers need not be liquid – they could be cognitive, imaging, functional, physiological, or other. They also need not be single or simple. They can be derived from numerous individual measurements across multiple dimensions. But they must be connected to causality, mechanism, or function—or they might be judged solely on Bonferroni-corrected p-values and will be disregarded. Use of these biomarkers will improve epistemology and lead to identification of mechanisms and relative contributions in populations and individuals.

### Systems cannot be understood a single dimension at a time

A recurring criticism of multimodal trials is that they should never be undertaken until all single components of the multimodal intervention have been previously tested individually in a portfolio of many unimodal intervention trials. However, this is not possible. If near-infinite resources, volunteers, and time were available—and if the number of possible interventions were finite—one could conduct a clinical trial on every intervention in isolation, then combine those that worked additively to achieve the optimal multidomain intervention. However, some interventions have infinite variations (e.g., Yang et al., 2023), including diet and exercise (considering all doses, frequencies, and modalities): multimodal trials are inevitable and necessary even with improving technology and trial design.

Furthermore, we expect interventions to be synergistic, not additive. Therefore, a combinatoric approach to a clinical trial portfolio is not possible. Notably—as we have previously discussed (Roach, Hodes, et al., 2022)— such a research portfolio would take centuries, cost too much, and require more participants than are recruitable. In practice, embracing such a portfolio would mean no sufficiently complex multimodal intervention would ever be tested.

We designed the COCOA and its PREVENTION trials to address these concerns (e.g., Roach, Hara, Fridman, et al., 2022; McEwen et al., 2021). Soon, it may be possible to test many thousands of hypotheses in a single clinical trial using modern high-throughput technologies and clever epistemology—COCOA can be considered a step in that direction. Indeed, the practice of designing trials that look at overarching “patterns” of diet or other behavioral interventions — such as the Dietary Approaches to Stop Hypertension (DASH) trial (Appel et al., 1997) — has gotten wider acceptance in recent years. Like DASH, COCOA is not specifically designed to be a parallel test of many individual components but a synergistic test of a multimodal intervention that differs in particulars between participants due to personalization of the intervention. Collection of dense data enables systems quantification. This systems quantification enables improvement of recommendations for multimodal interventions by identifying molecular mediators and/or biomarkers that reflect mechanisms that may identify and explain the effects of individual components of the interventions that are most beneficial in particular individuals (Bougea & Gourzis, 2024; Mielke et al., 2024).

### Dense data enables clinical trials to perceive a whole greater than the sum of parts

Investigation of a complex system by perturbation of a single variable at a time is slow and will never provide more than marginal understanding. Classical clinical trials yield only marginal distributions of outcomes. The knowledge they provide is akin to what can be learned from querying a laptop computer with an ohmmeter. Many drugs, such as lecanemab, will continue to be identified with such classic FDA-sanctioned approaches. Nevertheless, given that the best therapies for AD are likely to be multidomain, newer epistemologies will be required to understand them sufficiently to fully realize their potential. Although we agree with a recent editorial that more multimodal RCTs are needed (“Dementia prevention needs clinical trials”, 2025), these RCTs need to be carefully designed and used in conjunction with studies that generate knowledge of mechanism.

### Primary outcome measures

No pair of RCTs listed in **Table 2** share any primary outcome measure. On the one hand, this speaks to the robustness of the benefits observed from multidomain interventions, as most of the trials showed benefit. It also underscores the evolving nature of outcome measures, and the continued quest to find better measures. On the other hand, it makes it difficult to compare trials or to perform integrative analyses (e.g., validation or meta-analyses) based on these primary outcome measures. Many trials report secondary measures or multiomics datasets, which do enable such integrative analyses. Insofar as Cohen’s d is a valued measure of effect size, then increase in accuracy and precision of the outcome measure (e.g., MPI Score vs composite Z-scores) may increase Cohen’s d (because of the effect of measurement error, or standard deviation, on Cohen’s d), resulting in improved sense of importance of trial results (see also **Supplemental Figures 2-4**). Among trial results (**Table 2**), there is considerable variation in the statistical parameters (Cohen’s d and p-value) that are dependent on the accuracy and precision of the primary outcome measure. The trial with the most precise outcome measure (MPI score for COCOA) has the most significance but does not necessarily show a clinical effect (e.g., relative difference) greater than the other trials. One advantage of composite scores, such as bespoke composite scores or the more broadly accepted Clinical Dementia Rating – Sum of Boxes (CDR-SB), is they capture many dimensions of cognition in a single score; although the significance of the results from trials that use these as primary outcome measures may be lower, these composite scores suggest broad benefits of multidomain interventions across numerous cognitive dimensions. Despite that consideration, we recommend for future trials precise and accurate primary outcome measures to maximize power. In most cases, one cognitive and one functional primary outcome measure are warranted (McLaughlin et al., 2024). We recommend assigning composite measures as secondary outcomes to enable insights into robustness of results without sacrificing power.

### Insights

Lifestyle treatments are likely to complement, not replace, pharmaceutical treatments. Both are likely to be part of many optimal personalized multidomain therapies. Current pharmaceutical options have uncertain evidence (Osborne et al., 2023), limited effect duration, constrained target population, or side effects (e.g., Atwood & Perry, 2023), and offer limited range of targetable domains (e.g., most focus on proteostasis and produce marginal clinically relevant benefit as unimodal interventions). This presents an opportunity to develop a wider range of pharmaceuticals that target a range of treatment modes, and that can be easily adopted into personalized multidomain treatments. This need for multimodal therapies drives a need for cheaper drugs (cheaper to find, test, & make), that can be used in combination with each other to affect multiple systems simultaneously, and that allow for personalization (high safety and high ability to predict safety of drug combinations). Each individual drug might be used in only a small percentage of AD cases.

Lifestyle treatments tend to benefit even individuals without disease, so lifestyle interventions should be the foundation of multidomain therapies. Lifestyle interventions should be emphasized in current treatment for all individuals on the Alzheimer’s disease and related disorder (ADRD) spectrum. Together, the trials in **Table 2** provide strong evidence to elevate the prominence of these recommendations in guidelines and provider training. Lifestyle recommendations are often buried as afterthoughts at the end of multipage clinical recommendations that otherwise focus on pharmaceuticals (e.g., Alzheimer’s Association, 2019). Lifestyle recommendations often lack specifics, making them hard to implement, particularly for practitioners who are not experts. Strong recommendations for multimodal lifestyle intervention should be placed before pharmaceutical recommendations in professional clinical guidelines and given equal or greater amounts of text. More research — some of which is ongoing, such as WW-FINGERS (Kivipelto et al., 2020), including the Alzheimer’s Association U.S. Study to Protect Brain Health Through Lifestyle Intervention to Reduce Risk (U.S. POINTER) — is necessary to make this case even more convincing (Coon & Gómez-Morales, 2022).

More funding for lifestyle interventions should be applied both to research and via payers for medical care. The PREVENTION trial is nearing completion and should serve to iterate, validate, refine, and extend conclusions from COCOA and further test the methodologies discussed here (McEwen et al., 2021). Finally, pathways to *treat* ADRD may be different than pathways that *cause* ADRD (**Figure 1C**). Research should be pursued to identify methods of perturbing systems to benefit AD patients, even if the “opposite” of those system perturbations would not cause AD. Hysteresis is a common property of systems, and there may be multiple paths into and out of particular attractor states; not all of these paths are reversible.

### A plea for larger trials is hollow

Many papers end with an entreaty for larger and longer clinical trials. The motivation for such entreaties is that large trials are needed to generate small p-values and greater significance. The downside of such entreaties is that large trials that address all epistemological needs for AD translational research are either absolutely impossible (neither enough enrollable patients nor time in the universe) or practically impossible (no political appetite or commercial incentive to fund such trials). Furthermore, heterogeneity in large trials may do more to decrease than to increase power. Certainly, large trials should be encouraged and advocated for, and can address vital questions, particularly in large health care settings, such as whether particular policy implementations statistically improve population outcomes and save health care system dollars. Even though much of the basic knowledge necessary to drive rational implementation of multidomain intervention policies is already known, more knowledge is necessary to refine and improve interventions and broaden access to multidomain care. Such knowledge will have to come mostly from modern analyses of smaller cohorts and trials, given the impossibilities of all but a few large trials.

### Limitations of this scoping review

The primary limitation of this scoping review—and a major limitation of AD research—is the lack of clear diagnostic criteria for AD, particularly in presymptomatic stages. There are many trials for prevention of cognitive decline in healthy individuals. Unless a preponderance of these “healthy” individuals has presymptomatic AD, such trials would be excluded. However, few published studies on healthy individuals report the percentage of participants with presymptomatic AD. This presumably would require either sophisticated population-level epidemiologic inferences and/or expensive molecular or imaging biomarkers, many of which were unreliable at the time these trials were designed. Therefore, inclusion or exclusion of trials at FAST stage 1 may be error prone. Estimation of values not explicitly reported in published reports (including age at enrollment, FAST stage, and effect sizes) may be error prone. Trials not referenced in the literature or indexed in PubMed or ClinicalTrials.gov may have been missed from this compilation. Trials not including key search terms, such as “multimodal” or “multidomain” may have been missed.

### Recommended Policy and Clinical implications

We need to educate adults about options to prevent and treat cognitive decline and dementia. Public health policy should include elements to address modifiable risk factors for cognitive decline and ADRD. In clinical trials, better assessments for outcome measurements are needed, including those that precisely and accurately measure specific domains of cognition and function; imprecise aggregate measures should be reserved for secondary outcome measures. Systems quantification should be universally deployed in trials, and more widely utilized to guide personalized medicine and health. Personalized coaching in a variety of forms (e.g., individual, group, AI-assisted) will be valuable to many; payors should support such coaching. Very inexpensive population-level options for each domain should be developed and deployed using advances in remote medicine to increase the reach of care to all individuals. The American Academy of Neurology’s (AAN) Committee on Public Engagement position statement on national brain plans (Rost et al., 2023) includes calls to (1) optimize brain health through integration of preventive care practices, and (2) include brain health checkups as part of the standard of care across the lifespan. Switzerland has such a national plan that incorporates many of these ideas and recognizes the utility and need for multidomain interventions delaying age-associated cognitive deterioration (Bassetti et al., 2023), alongside blood biomarkers for population screening, and including effective pharmaceuticals to decrease the incidence of cognitive impairment and dementia. All nations should follow this lead and implement national brain health plans.

Multidomain interventions are best tailored to the individual’s personal circumstances (including genetic, environmental, and sociocultural context). Therefore, the best way to advance knowledge for any complex disease is a research program spanning many studies, many groups, and many years. However, most simple clinical trial designs would not be part of this research program, as they would provide only a narrow glimpse of the systems landscape. Each human differs from all others across thousands if not millions or billions of parameters. Efforts to create uniformity at baseline by matching on a few—or even a few dozen—of these parameters, such as race or sex or education, have some value but at best reduce the risk of confounding by a little while decreasing diversity a lot. Not only does a decrease in diversity impair society, but it also limits the generalizability and validity of translatable knowledge resulting from a trial.

Differences between participants are a strength as well as a limitation of all clinical trials. Although analytical methods for systems biology vary widely—ranging from statistical learning to mechanistic network modeling— they all require diverse perturbations on diverse systems. Classic trials designed to test the marginal effect of a single variable emphasize constancy on all other variables (“everything else being equal”) — particularly those that could influence the outcome. Constancy avoids confounding—the possibility that causal influences on the outcome are not due to the manipulated variable of interest, but rather to other variable(s) that exhibit diversity either at baseline or subsequently. But—in the context of complex diseases—such trial design has always been a mirage. Everything else is not equal: humans have far more variability than can be measured, let alone be controlled or be held constant. Of this variability, sex is particularly important, as the causes of AD differ in strength between men and women, and the response to lifestyle interventions differ (Zülke et al., 2023). The nature of complex disease is such that many variables that affect the outcome are likely to be unanticipated. Therefore, it is best to measure the entire system of each individual, understand them deeply, and model their trajectories. Biological plausibility must anchor epistemology (Roach & Freidin, 2023; Uleman et al., 2024).

This is hard—particularly with twentieth century analytical techniques—but must be achieved if we are to make substantial progress in understanding and treating complex diseases in this century. In the immediate future, we recommend AD clinical studies trials be based on observational deep data of ∼1000 patients. Such studies will capture data from individuals transitioning through different subgroups and different stages of AD, maximizing their value for systems biological analysis.

In this context of maximizing diversity by broadening inclusion and exclusion criteria, there is also value in focusing on particular populations. These populations should include individuals: (1) likely to have AD— allowing efficient allocation of therapeutic resources, (2) at earlier stages of pathogenesis more likely to be interruptible or reversible than later stages (Aisen et al., 2022), and (3) with early-stage AD with sufficient function to understand and respond to coaching to implement multidomain recommendations. Furthermore, solely from a perspective of trial design—to have statistical power and to complete the trial in a practical timespan—we desire to see a large and significant effect of the intervention over a short period of time. In early stages of the AD spectrum, control trajectories are likely to decline very slowly if at all, and a very large and/or long trial might be required. In dementia stages, individuals may be too impaired to respond to telephonically coached multidomain interventions. Therefore, pilot trials should focus on individuals partially along the AD spectrum. Ultimately, a priority for policymakers should be to enable multidomain interventions that can be conducted very inexpensively and burden free, so that they can be targeted at early stages of disease to entire populations. Improved early detection and risk stratification should fine-tune and target these interventions.

## Contributions

All authors participated in all aspects of manuscript preparation. The authors have no acknowledgments to report. The authors have no conflict of interest to report. All authors have seen and approved the manuscript.

## Ethics and Reporting

All relevant ethical guidelines have been followed. As this is a review article, none of the following were necessary for the writing of this manuscript: IRB and/or ethics committee approvals, patient/participant consent, or appropriate institutional forms archived. This information, as well as clinical trial IDs, can be found in the referenced papers in which each clinical trial was reported.

## Data Availability

No new data were generated for this manuscript.

## Supporting information

PRISMA Checklist

## REFERENCES

Abramowitz A, Weber M. Management of MCI in the Outpatient Setting. Curr Psychiatry Rep. 2024 Aug;26(8):413–421. doi: 10.1007/s11920-024-01514-3. Epub 2024 Jun 10. PMID: 38856858.

Aihara K, Liu R, Koizumi K, Liu X, Chen L. Dynamical network biomarkers: Theory and applications. Gene. 2022 Jan 15;808:145997. doi: 10.1016/j.gene.2021.145997. Epub 2021 Oct 6. PMID: 34626720.

Aisen PS, Jimenez-Maggiora GA, Rafii MS, Walter S, Raman R. Early-stage Alzheimer disease: getting trial-ready. Nat Rev Neurol. 2022 Jul;18(7):389–399. doi: 10.1038/s41582-022-00645-6. Epub 2022 Apr 4. PMID: 35379951; PMCID: PMC8978175.

Alzheimer’s Association. Basic Resources on Alzheimer’s for Primary Care Physicians. Alzheimer’s Association Green-Field Library. 2019.

Alzheimer’s Network for Treatment and Diagnostics (ALZ-NET). ALZ-NET Protocol. Version: January 10, 2025.

Amini Y, Saif N, Greer C, Hristov H, Isaacson R. The Role of Nutrition in Individualized Alzheimer’s Risk Reduction. Curr Nutr Rep. 2020 Jun;9(2):55–63. doi: 10.1007/s13668-020-00311-7. PMID: 32277428.

Andrieu S, Guyonnet S, Coley N, Cantet C, Bonnefoy M, Bordes S, Bories L, Cufi MN, Dantoine T, Dartigues JF, Desclaux F, Gabelle A, Gasnier Y, Pesce A, Sudres K, Touchon J, Robert P, Rouaud O, Legrand P, Payoux P, Caubere JP, Weiner M, Carrié I, Ousset PJ, Vellas B; MAPT Study Group. Effect of long-term omega 3 polyunsaturated fatty acid supplementation with or without multidomain intervention on cognitive function in elderly adults with memory complaints (MAPT): a randomised, placebo-controlled trial. Lancet Neurol. 2017 May;16(5):377–389. doi: 10.1016/S1474-4422(17)30040-6. Epub 2017 Mar 27. PMID: 28359749.

Appel LJ, Moore TJ, Obarzanek E, Vollmer WM, Svetkey LP, Sacks FM, Bray GA, Vogt TM, Cutler JA, Windhauser MM, Lin PH, Karanja N. A clinical trial of the effects of dietary patterns on blood pressure. DASH Collaborative Research Group. N Engl J Med. 1997 Apr 17;336(16):1117–24. doi: 10.1056/NEJM199704173361601. PMID: 9099655.

Arnold M, Nho K, Kueider-Paisley A, Massaro T, Huynh K, Brauner B, MahmoudianDehkordi S, Louie G, Moseley MA, Thompson JW, John-Williams LS, Tenenbaum JD, Blach C, Chang R, Brinton RD, Baillie R, Han X, Trojanowski JQ, Shaw LM, Martins R, Weiner MW, Trushina E, Toledo JB, Meikle PJ, Bennett DA, Krumsiek J, Doraiswamy PM, Saykin AJ, Kaddurah-Daouk R, Kastenmüller G. Sex and APOE ε4 genotype modify the Alzheimer’s disease serum metabolome. Nat Commun. 2020 Mar 2;11(1):1148. doi: 10.1038/s41467-020-14959-w. PMID: 32123170; PMCID: PMC7052223.

Atwood CS, Perry G. Playing Russian Roulette with Alzheimer’s Disease Patients: Do the Cognitive Benefits of Lecanemab Outweigh the Risk of Edema, Stroke and Encephalitis? J Alzheimers Dis. 2023;92(3):799–801. doi: 10.3233/JAD-230040. PMID: 36847013.

Bae S, Lee S, Lee S, Jung S, Makino K, Harada K, Harada K, Shinkai Y, Chiba I, Shimada H. The effect of a multicomponent intervention to promote community activity on cognitive function in older adults with mild cognitive impairment: A randomized controlled trial. Complement Ther Med. 2019 Feb;42:164–169. doi: 10.1016/j.ctim.2018.11.011. Epub 2018 Nov 13. PMID: 30670238.

Baker LD, Snyder HM, Espeland MA, Whitmer RA, Kivipelto M, Woolard N, Katula J, Papp KV, Ventrelle J, Graef S, Hill MA, Rushing S, Spell J, Lovato L, Felton D, Williams BJ, Ghadimi Nouran M, Raman R, Ngandu T, Solomon A, Wilmoth S, Cleveland ML, Williamson JD, Lambert KL, Tomaszewski Farias S, Day CE, Tangney CC, Gitelman DR, Matongo O, Reynolds T, Pavlik VN, Yu MM, Alexander AS, Elbein R, McDonald AM, Salloway S, Wing RR, Antkowiak S, Morris MC, Carrillo MC; U.S. POINTER Study Group. Study design and methods: U.S. study to protect brain health through lifestyle intervention to reduce risk (U.S. POINTER). Alzheimers Dement. 2024 Feb;20(2):769–782. doi: 10.1002/alz.13365. Epub 2023 Sep 30. PMID: 37776210; PMCID: PMC10916955.

Bassetti CLA, Heldner MR, Adorjan K, Albanese E, Allali G, Arnold M, Bègue I, Bochud M, Chan A, do Cuénod KQ, et al. The Swiss Brain Health Plan 2023–2033. Clinical and Translational Neuroscience. 2023; 7(4):38. 10.3390/ctn7040038.

Bermejo-Pareja F, Del Ser T. Controversial Past, Splendid Present, Unpredictable Future: A Brief Review of Alzheimer Disease History. J Clin Med. 2024 Jan 17;13(2):536. doi: 10.3390/jcm13020536. PMID: 38256670; PMCID: PMC10816332.

Birks JS, Harvey RJ. Donepezil for dementia due to Alzheimer’s disease. Cochrane Database Syst Rev. 2018 Jun 18;6(6):CD001190. doi: 10.1002/14651858.CD001190.pub3. PMID: 29923184; PMCID: PMC6513124.

Blumenthal JA, Smith PJ, Mabe S, Hinderliter A, Welsh-Bohmer K, Browndyke JN, Doraiswamy PM, Lin PH, Kraus WE, Burke JR, Sherwood A. Longer Term Effects of Diet and Exercise on Neurocognition: 1-Year Follow-up of the ENLIGHTEN Trial. J Am Geriatr Soc. 2020 Mar;68(3):559–568. doi: 10.1111/jgs.16252. Epub 2019 Nov 22. PMID: 31755550; PMCID: PMC7056586.

Brodaty H, Chau T, Heffernan M, Ginige JA, Andrews G, Millard M, Sachdev PS, Anstey KJ, Lautenschlager NT, McNeil JJ, Jorm L, Kochan NA, Maeder A, Welberry H, San Jose JC, Briggs NE, Popovic G, Mavros Y, Almendrales Rangel C, Noble Y, Radd-Vagenas S, Flood VM, O’Leary F, Lampit A, Walton CC, Barr P, Fiatarone Singh M, Valenzuela M. An online multidomain lifestyle intervention to prevent cognitive decline in at-risk older adults: a randomized controlled trial. Nat Med. 2025 Feb;31(2):565–573. doi: 10.1038/s41591-024-03351-6. Epub 2025 Jan 28. PMID: 39875685.

Bougea A, Gourzis P. Biomarker-Based Precision Therapy for Alzheimer’s Disease: Multidimensional Evidence Leading a New Breakthrough in Personalized Medicine. J Clin Med. 2024 Aug 8;13(16):4661. doi: 10.3390/jcm13164661. PMID: 39200803; PMCID: PMC11355840.

Chen LK, Hwang AC, Lee WJ, Peng LN, Lin MH, Neil DL, Shih SF, Loh CH, Chiou ST; Taiwan Health Promotion Intervention Study for Elders research group. Efficacy of multidomain interventions to improve physical frailty, depression and cognition: data from cluster-randomized controlled trials. J Cachexia Sarcopenia Muscle. 2020 Jun;11(3):650–662. doi: 10.1002/jcsm.12534. Epub 2020 Mar 5. PMID: 32134208; PMCID: PMC7296266.

Chen P, Li Y, Liu X, Liu R, Chen L. Detecting the tipping points in a three-state model of complex diseases by temporal differential networks. J Transl Med. 2017 Oct 26;15(1):217. doi: 10.1186/s12967-017-1320-7. PMID: 29073904; PMCID: PMC5658963.

Chhetri JK, de Souto Barreto P, Cantet C, Pothier K, Cesari M, Andrieu S, Coley N, Vellas B. Effects of a 3-Year Multi-Domain Intervention with or without Omega-3 Supplementation on Cognitive Functions in Older Subjects with Increased CAIDE Dementia Scores. J Alzheimers Dis. 2018;64(1):71–78. doi: 10.3233/JAD-180209. PMID: 29865075.

Coon DW, Gómez-Morales A. Modifiable Risk Factors for Brain Health and Dementia and Opportunities for Intervention: A Brief Review. Clin Gerontol. 2022 Aug 22:1–12. doi: 10.1080/07317115.2022.2114396. Epub ahead of print. PMID: 35996225.

Damirchi A, Hosseini F, Babaei P. Mental Training Enhances Cognitive Function and BDNF More Than Either Physical or Combined Training in Elderly Women With MCI: A Small-Scale Study. Am J Alzheimers Dis Other Demen. 2018 Feb;33(1):20–29. doi: 10.1177/1533317517727068. Epub 2017 Sep 25. PMID: 28946752; PMCID: PMC10852433.

Delport A, Hewer R. The amyloid precursor protein: a converging point in Alzheimer’s disease. Mol Neurobiol. 2022 Jul;59(7):4501–4516. doi: 10.1007/s12035-022-02863-x. Epub 2022 May 17. PMID: 35579846.

Dementia prevention needs clinical trials. Nat Med. 2025 Feb;31(2):353. doi: 10.1038/s41591-025-03552-7. PMID: 39972238.

Diener C, Holscher HD, Filek K, Corbin KD, Moissl-Eichinger C, Gibbons SM. Metagenomic estimation of dietary intake from human stool. Nat Metab. 2025 Mar;7(3):617–630. doi: 10.1038/s42255-025-01220-1. Epub 2025 Feb 18. Erratum in: Nat Metab. 2025 Mar;7(3):633. doi: 10.1038/s42255-025-01284-z. PMID: 39966520; PMCID: PMC11949708.

Dubois B, Villain N, Schneider L, Fox N, Campbell N, Galasko D, Kivipelto M, Jessen F, Hanseeuw B, Boada M, Barkhof F, Nordberg A, Froelich L, Waldemar G, Frederiksen KS, Padovani A, Planche V, Rowe C, Bejanin A, Ibanez A, Cappa S, Caramelli P, Nitrini R, Allegri R, Slachevsky A, de Souza LC, Bozoki A, Widera E, Blennow K, Ritchie C, Agronin M, Lopera F, Delano-Wood L, Bombois S, Levy R, Thambisetty M, Georges J, Jones DT, Lavretsky H, Schott J, Gatchel J, Swantek S, Newhouse P, Feldman HH, Frisoni GB. Alzheimer Disease as a Clinical-Biological Construct-An International Working Group Recommendation. JAMA Neurol. 2024 Dec 1;81(12):1304–1311. doi: 10.1001/jamaneurol.2024.3770. PMID: 39483064.

Espay AJ, Lang AE. Parkinson Diseases in the 2020s and Beyond: Replacing Clinico-Pathologic Convergence With Systems Biology Divergence. J Parkinsons Dis. 2018;8(s1):S59-S64. doi: 10.3233/JPD-181465. PMID: 30584155; PMCID: PMC6311362.

Fan M, Li Q, Yang T, Yang Y, Chen Z, Xuan G, Ruan Y, Sun S, Wang M, Chen X, Huang Y, Yang Z, Wang Y. Effect of Multimodal Intervention in Individuals with Mild Cognitive Impairment: A Randomized Clinical Trial in Shanghai. J Alzheimers Dis. 2024;101(1):235–248. doi: 10.3233/JAD-231370. PMID: 39031354; PMCID: PMC11380217.

Fortea J, Pegueroles J, Alcolea D, Belbin O, Dols-Icardo O, Vaqué-Alcázar L, Videla L, Gispert JD, Suárez-Calvet M, Johnson SC, Sperling R, Bejanin A, Lleó A, Montal V. APOE4 homozygozity represents a distinct genetic form of Alzheimer’s disease. Nat Med. 2024 May;30(5):1284–1291. doi: 10.1038/s41591-024-02931-w. Epub 2024 May 6. Erratum in: Nat Med. 2024 Jul;30(7):2093. doi: 10.1038/s41591-024-03127-y. PMID: 38710950.

Fedak KM, Bernal A, Capshaw ZA, Gross S. Applying the Bradford Hill criteria in the 21st century: how data integration has changed causal inference in molecular epidemiology. Emerg Themes Epidemiol. 2015;12:14. Published 2015 Sep 30. doi:10.1186/s12982-015-0037-4.

Ferrer I. Hypothesis review: Alzheimer’s overture guidelines. Brain Pathol. 2023 Jan;33(1):e13122. doi: 10.1111/bpa.13122. Epub 2022 Oct 12. PMID: 36223647; PMCID: PMC9836379.

George DR, Qualls SH, Camp CJ, Whitehouse PJ. Renovating Alzheimer’s: “constructive” reflections on the new clinical and research diagnostic guidelines. Gerontologist. 2013 Jun;53(3):378–87. doi: 10.1093/geront/gns096. Epub 2012 Aug 30. PMID: 22936533.

Gong CX, Dai CL, Liu F, Iqbal K. Multi-Targets: An Unconventional Drug Development Strategy for Alzheimer’s Disease. Front Aging Neurosci. 2022 Feb 9;14:837649. doi: 10.3389/fnagi.2022.837649. PMID: 35222001; PMCID: PMC8864545.

Higginbotham L, Carter EK, Dammer EB, Haque RU, Johnson ECB, Duong DM, Yin L, De Jager PL, Bennett DA, Felsky D, Tio ES, Lah JJ, Levey AI, Seyfried NT. Unbiased classification of the elderly human brain proteome resolves distinct clinical and pathophysiological subtypes of cognitive impairment. Neurobiol Dis. 2023 Oct 1;186:106286. doi: 10.1016/j.nbd.2023.106286. Epub 2023 Sep 7. PMID: 37689213; PMCID: PMC10750427.

Huang Y, Mucke L. Alzheimer mechanisms and therapeutic strategies. Cell. 2012 Mar 16;148(6):1204-22. doi: 10.1016/j.cell.2012.02.040. PMID: 22424230; PMCID: PMC3319071.

Isaacson RS, Hristov H, Saif N, Hackett K, Hendrix S, Melendez J, Safdieh J, Fink M, Thambisetty M, Sadek G, Bellara S, Lee P, Berkowitz C, Rahman A, Meléndez-Cabrero J, Caesar E, Cohen R, Lu PL, Dickson SP, Hwang MJ, Scheyer O, Mureb M, Schelke MW, Niotis K, Greer CE, Attia P, Mosconi L, Krikorian R. Individualized clinical management of patients at risk for Alzheimer’s dementia. Alzheimers Dement. 2019 Dec;15(12):1588–1602. doi: 10.1016/j.jalz.2019.08.198. Epub 2019 Oct 31. PMID: 31677936; PMCID: PMC6925647.

Jagust WJ. The changing definition of Alzheimer’s disease. Lancet Neurol. 2021 Jun;20(6):414–415. doi: 10.1016/S1474-4422(21)00077-6. Epub 2021 Apr 29. PMID: 33933185.

Kaufman JS, Poole C. Looking back on “causal thinking in the health sciences”. Annu Rev Public Health. 2000;21:101–19. doi: 10.1146/annurev.publhealth.21.1.101. PMID: 10884948.

Kikuchi M, Kobayashi K, Itoh S, Kasuga K, Miyashita A, Ikeuchi T, Yumoto E, Kosaka Y, Fushimi Y, Takeda T, Manabe S, Hattori S; Alzheimer’s Disease Neuroimaging Initiative; Nakaya A, Kamijo K, Matsumura Y. Identification of mild cognitive impairment subtypes predicting conversion to Alzheimer’s disease using multimodal data. Comput Struct Biotechnol J. 2022 Aug 22;20:5296–5308. doi: 10.1016/j.csbj.2022.08.007. PMID: 36212530; PMCID: PMC9513733.

Kivipelto M, Mangialasche F, Snyder HM, Allegri R, Andrieu S, Arai H, Baker L, Belleville S, Brodaty H, Brucki SM, Calandri I, Caramelli P, Chen C, Chertkow H, Chew E, Choi SH, Chowdhary N, Crivelli L, Torre R, Du Y, Dua T, Espeland M, Feldman HH, Hartmanis M, Hartmann T, Heffernan M, Henry CJ, Hong CH, Håkansson K, Iwatsubo T, Jeong JH, Jimenez-Maggiora G, Koo EH, Launer LJ, Lehtisalo J, Lopera F, Martínez-Lage P, Martins R, Middleton L, Molinuevo JL, Montero-Odasso M, Moon SY, Morales-Pérez K, Nitrini R, Nygaard HB, Park YK, Peltonen M, Qiu C, Quiroz YT, Raman R, Rao N, Ravindranath V, Rosenberg A, Sakurai T, Salinas RM, Scheltens P, Sevlever G, Soininen H, Sosa AL, Suemoto CK, Tainta-Cuezva M, Velilla L, Wang Y, Whitmer R, Xu X, Bain LJ, Solomon A, Ngandu T, Carrillo MC. World-Wide FINGERS Network: A global approach to risk reduction and prevention of dementia. Alzheimers Dement. 2020 Jul;16(7):1078–1094. doi: 10.1002/alz.12123. Epub 2020 Jul 5. PMID: 32627328; PMCID: PMC9527644.

Knopman DS, Petersen RC, Jack CR Jr. A brief history of “Alzheimer disease”: Multiple meanings separated by a common name. Neurology. 2019 May 28;92(22):1053–1059. doi: 10.1212/WNL.0000000000007583. Epub 2019 Apr 26. PMID: 31028129; PMCID: PMC6556090.

Krieger MM, Richter RR, Austin TM. An exploratory analysis of PubMed’s free full-text limit on citation retrieval for clinical questions. J Med Libr Assoc. 2008 Oct;96(4):351–5. doi: 10.3163/1536-5050.96.4.010. PMID: 18974812; PMCID: PMC2568849.

Lanctôt KL, Boada M, Tariot PN, Dabbous F, Hahn-Pedersen J, Udayachalerm S, Raket LL, Saiontz-Martinez C, Michalak W, Weidner W, Cummings J. Association between clinical dementia rating and clinical outcomes in Alzheimer’s disease. Alzheimers Dement (Amst). 2024 Jan 17;16(1):e12522. doi: 10.1002/dad2.12522. PMID: 38239329; PMCID: PMC10794857.

Lin J, Dong B, Vellas B. Editorial: Preventive Trials for Alzheimer’s Diseases: The Multi-domain and the Targeted Therapies Approaches Will Have to Be Associated. J Nutr Health Aging. 2016;20(5):494–5. doi: 10.1007/s12603-016-0724-z. PMID: 27102785.

Liu R, Chen P, Chen L. Single-sample landscape entropy reveals the imminent phase transition during disease progression. Bioinformatics. 2020 Mar 1;36(5):1522–1532. doi: 10.1093/bioinformatics/btz758. Erratum in: Bioinformatics. 2020 Apr 15;36(8):2644. PMID: 31598632.

Livingston G, Huntley J, Liu KY, Costafreda SG, Selbæk G, Alladi S, Ames D, Banerjee S, Burns A, Brayne C, Fox NC, Ferri CP, Gitlin LN, Howard R, Kales HC, Kivimäki M, Larson EB, Nakasujja N, Rockwood K, Samus Q, Shirai K, Singh-Manoux A, Schneider LS, Walsh S, Yao Y, Sommerlad A, Mukadam N. Dementia prevention, intervention, and care: 2024 report of the Lancet standing Commission. Lancet. 2024 Aug 10;404(10452):572-628. doi: 10.1016/S0140-6736(24)01296-0. Epub 2024 Jul 31. PMID: 39096926.)

Livingston G, Huntley J, Sommerlad A, Ames D, Ballard C, Banerjee S, Brayne C, Burns A, Cohen-Mansfield J, Cooper C, Costafreda SG, Dias A, Fox N, Gitlin LN, Howard R, Kales HC, Kivimäki M, Larson EB, Ogunniyi A, Orgeta V, Ritchie K, Rockwood K, Sampson EL, Samus Q, Schneider LS, Selbæk G, Teri L, Mukadam N. Dementia prevention, intervention, and care: 2020 report of the Lancet Commission. Lancet. 2020 Aug 8;396(10248):413-446. doi: 10.1016/S0140-6736(20)30367-6. Epub 2020 Jul 30. PMID: 32738937; PMCID: PMC7392084.

McEwen SC, Merrill DA, Bramen J, Porter V, Panos S, Kaiser S, Hodes J, Ganapathi A, Bell L, Bookheimer T, Glatt R, Rapozo M, Ross MK, Price ND, Kelly D, Funk CC, Hood L, Roach JC. A systems-biology clinical trial of a personalized multimodal lifestyle intervention for early Alzheimer’s disease. Alzheimers Dement (N Y). 2021 Jul 20;7(1):e12191. doi: 10.1002/trc2.12191. PMID: 34295960; PMCID: PMC8290633.

Masurkar AV, Marsh K, Morgan B, Leitner D, Wisniewski T. Factors Affecting Resilience and Prevention of Alzheimer’s Disease and Related Dementias. Ann Neurol. 2024 Oct;96(4):633–649. doi: 10.1002/ana.27055. Epub 2024 Aug 17. PMID: 39152774; PMCID: PMC11534551.

McLaughlin J, Scotton WJ, Ryan NS, Hardy JA, Shoai M. Assessing clinical progression measures in Alzheimer’s disease trials: A systematic review and meta-analysis. Alzheimers Dement. 2024 Dec;20(12):8673–8683. doi: 10.1002/alz.14314. Epub 2024 Oct 22. PMID: 39439251; PMCID: PMC11667530.

Mielke MM, Anderson M, Ashford JW, Jeromin A, Lin PJ, Rosen A, Tyrone J, Vandevrede L, Willis DR, Hansson O, Khachaturian AS, Schindler SE, Weiss J, Batrla R, Bozeat S, Dwyer JR, Holzapfel D, Jones DR, Murray JF, Partrick KA, Scholler E, Vradenburg G, Young D, Braunstein JB, Burnham SC, de Oliveira FF, Hu YH, Mattke S, Merali Z, Monane M, Sabbagh MN, Shobin E, Weiner M, Udeh-Momoh CT. Recommendations for clinical implementation of blood-based biomarkers for Alzheimer’s disease. Alzheimers Dement. 2024 Nov;20(11):8216–8224. doi: 10.1002/alz.14184. Epub 2024 Oct 1. PMID: 39351838; PMCID: PMC11567872.

Neff RA, Wang M, Vatansever S, Guo L, Ming C, Wang Q, Wang E, Horgusluoglu-Moloch E, Song WM, Li A, Castranio EL, Tcw J, Ho L, Goate A, Fossati V, Noggle S, Gandy S, Ehrlich ME, Katsel P, Schadt E, Cai D, Brennand KJ, Haroutunian V, Zhang B. Molecular subtyping of Alzheimer’s disease using RNA sequencing data reveals novel mechanisms and targets. Sci Adv. 2021 Jan 6;7(2):eabb5398. doi: 10.1126/sciadv.abb5398. PMID: 33523961; PMCID: PMC7787497.

Ngandu T, Lehtisalo J, Korkki S, Solomon A, Coley N, Antikainen R, Bäckman L, Hänninen T, Lindström J, Laatikainen T, Paajanen T, Havulinna S, Peltonen M, Neely AS, Strandberg T, Tuomilehto J, Soininen H, Kivipelto M. The effect of adherence on cognition in a multidomain lifestyle intervention (FINGER). Alzheimers Dement. 2022 Jul;18(7):1325–1334. doi: 10.1002/alz.12492. Epub 2021 Oct 20. PMID: 34668644.

Niotis K, Janney C, Helfman S, Hristov H, Clute-Reinig N, Angerbauer D, Saperia C, Murray S, Westine J, Seifan A, Melendez-Herencia J, Parthasarathy P, Colvee H, Lewis B, Lakis J, Sisser P, Dishary J, Mosse M, Saville D, Rumberger A, McCullough M, Isaacson R. A Blood Biomarker-guided Precision Medicine Approach for Individualized Neurodegenerative Disease Risk Reduction and Treatment: The Future of Preventive Neurology? (P7-3.016). Neurology. 2025 Apr 8;104(7_Supplement_1):S7-3.016. doi: 10.1212/WNL.0000000000208443.

Niotis K, Saperia C, Saif N, Carlton C, Isaacson RS. Alzheimer’s disease risk reduction in clinical practice: a priority in the emerging field of preventive neurology. Nat Mental Health. 2024 Jan;2(1):25–40. doi:10.1038/s44220-023-00191-0.

Norton S, Matthews FE, Barnes DE, Yaffe K, Brayne C. Potential for primary prevention of Alzheimer’s disease: an analysis of population-based data. Lancet Neurol. 2014 Aug;13(8):788–94. doi: 10.1016/S1474-4422(14)70136-X. Erratum in: Lancet Neurol. 2014 Nov;13(11):1070. PMID: 25030513.

Norwitz NG, Saif N, Ariza IE, Isaacson RS. Precision Nutrition for Alzheimer’s Prevention in ApoE4 Carriers. Nutrients. 2021 Apr 19;13(4):1362. doi: 10.3390/nu13041362. PMID: 33921683; PMCID: PMC8073598.

Ornish, D., Madison, C., Kivipelto, M. et al. Effects of intensive lifestyle changes on the progression of mild cognitive impairment or early dementia due to Alzheimer’s disease: a randomized, controlled clinical trial. Alz Res Therapy 16, 122 (2024). 10.1186/s13195-024-01482-z.

Osborne OM, Naranjo O, Heckmann BL, Dykxhoorn D, Toborek M. Anti-amyloid: An antibody to cure Alzheimer’s or an attitude. iScience. 2023 Jul 24;26(8):107461. doi: 10.1016/j.isci.2023.107461. PMID: 37588168; PMCID: PMC10425904.

Paterson T, Jennifer Rohrs, Timothy J. Hohman, Mark Mapstone, Allan I. Levey, LeRoy Hood, Alzheimer’s Disease Neuroimaging Initiative, Cory C. Funk. Multi-omic ADNI CSF and plasma data integration identifies distinct metabolic transitions in disease progression in Alzheimer’s Disease. bioRxiv 2024.07.23.604835; 10.1101/2024.07.23.604835.

Peterson, D. and Keeley, J.W. (2015). Syndrome, Disorder, and Disease. In The Encyclopedia of Clinical Psychology (eds R.L. Cautin and S.O. Lilienfeld). 10.1002/9781118625392.wbecp154.

Petersen RC, Caracciolo B, Brayne C, Gauthier S, Jelic V, Fratiglioni L. Mild cognitive impairment: a concept in evolution. J Intern Med. 2014 Mar;275(3):214–28. doi: 10.1111/joim.12190. PMID: 24605806; PMCID: PMC3967548.

Petersen RC, Mormino E, Schneider JA. Alzheimer Disease—What’s in a Name? JAMA Neurol. 2024;81(12):1245–1246. doi:10.1001/jamaneurol.2024.3766.

Pontzer, H. (2021). Burn: New Research Blows the Lid Off How We Really Burn Calories, Stay Healthy, and Lose Weight. New York, NY: Avery, an imprint of Penguin Random House.

Reisberg, B., Jamil, I. A., Khan, S., Monteiro, I., Torossian, C., Ferris, S., Sabbagh, M., Gauthier, S., Auer, S., Shulman, M. B., Kluger, A., Franssen, E., & Wegiel, J. (2010). Staging Dementia. In M. T. Abou-Saleh, C. Katona, & A. Kumar (Eds.), Principles and Practice of Geriatric Psychiatry (3rd ed., pp. 162–169). John Wiley & Sons, Ltd.

Roach JC, Edens L, Markewych DR, Rapozo MK, Hara J, Glusman G, Funk C, Bramen J, Baloni P, Shankle WR, Hood L. A multimodal intervention for Alzheimer’s disease results in multifaceted systemic effects reflected in blood and ameliorates functional and cognitive outcomes. 2022. medRxiv. https://www.medrxiv.org/content/10.1101/2022.09.27.22280385v2.

Roach JC, Freidin MB. Editorial: Insights in human and medical genomics: 2022. Front Genet. 2023 Sep 25;14:1287894. doi: 10.3389/fgene.2023.1287894. PMID: 37818104; PMCID: PMC10561311.

Roach JC, Hara J, Fridman D, Lovejoy JC, Jade K, Heim L, Romansik R, Swietlikowski A, Phillips S, Rapozo MK, Shay MA, Fischer D, Funk C, Dill L, Brant-Zawadzki M, Hood L, Shankle WR. The Coaching for Cognition in Alzheimer’s (COCOA) trial: Study design. Alzheimers Dement (N Y). 2022 Jul 26;8(1):e12318. doi: 10.1002/trc2.12318. PMID: 35910672; PMCID: PMC9322829.

Roach JC, Hodes JF, Funk CC, Shankle WR, Merrill DA, Hood L, Bramen J. Dense data enables twenty-first century clinical trials. Alzheimers Dement (N Y). 2022 Jun 13;8(1):e12297. doi: 10.1002/trc2.12297. PMID: 35733645; PMCID: PMC9191823.

Roach JC, Rapozo MK, Hara J, Glusman G, Lovejoy J, Shankle WR, Hood L; COCOA Consortium. A Remotely Coached Multimodal Lifestyle Intervention for Alzheimer’s Disease Ameliorates Functional and Cognitive Outcomes. J Alzheimers Dis. 2023;96(2):591–607. doi: 10.3233/JAD-230403. PMID: 37840487.

Rosenberg A, Mangialasche F, Ngandu T, Solomon A, Kivipelto M. Multidomain Interventions to Prevent Cognitive Impairment, Alzheimer’s Disease, and Dementia: From FINGER to World-Wide FINGERS. J Prev Alzheimers Dis. 2020;7(1):29–36. doi: 10.14283/jpad.2019.41. PMID: 32010923; PMCID: PMC7222931.

Rosenberg A, Ngandu T, Rusanen M, Antikainen R, Bäckman L, Havulinna S, Hänninen T, Laatikainen T, Lehtisalo J, Levälahti E, Lindström J, Paajanen T, Peltonen M, Soininen H, Stigsdotter-Neely A, Strandberg T, Tuomilehto J, Solomon A, Kivipelto M. Multidomain lifestyle intervention benefits a large elderly population at risk for cognitive decline and dementia regardless of baseline characteristics: The FINGER trial. Alzheimers Dement. 2018 Mar;14(3):263–270. doi: 10.1016/j.jalz.2017.09.006. Epub 2017 Oct 19. PMID: 29055814.

Rosenberg A, Untersteiner H, Guazzarini AG, Bödenler M, Bruinsma J, Buchgraber-Schnalzer B, Colombo M, Crutzen R, Diaz A, Fotiadis DI, Hilberger H, Huber S, Kaartinen N, Kassiotis T, Kivipelto M, Lehtisalo J, Loukas VS, Lötjönen J, Pirani M, Thunborg C, Hanke S, Mangialasche F, Mecocci P, Stögmann E, Ngandu T; on behaf of the LETHE Consortium. A digitally supported multimodal lifestyle program to promote brain health among older adults (the LETHE randomized controlled feasibility trial): study design, progress, and first results. Alzheimers Res Ther. 2024 Nov 21;16(1):252. doi: 10.1186/s13195-024-01615-4. PMID: 39574193; PMCID: PMC11580696.

Ross JA, Dodel R. Preclinical CSF proteomic changes: a milestone in biomarker detection for autosomal dominant Alzheimer’s disease. Signal Transduct Target Ther. 2025 Jan 20;10(1):16. doi: 10.1038/s41392-024-02109-3. PMID: 39828719; PMCID: PMC11743781.

Rost NS, Salinas J, Jordan JT, Banwell B, Correa DJ, Said RR, Selwa LM, Song S, Evans DA; American Academy of Neurology’s Committee on Public Engagement. The Brain Health Imperative in the 21st Century-A Call to Action: The AAN Brain Health Platform and Position Statement. Neurology. 2023 Sep 26;101(13):570-579. doi: 10.1212/WNL.0000000000207739. PMID: 37730439; PMCID: PMC10558159.

Rothman KJ. Causes. 1976. Am J Epidemiol. 1995 Jan 15;141(2):90-5; discussion 89. doi: 10.1093/oxfordjournals.aje.a117417. PMID: 7817976.

Saif N, Hristov H, Akiyoshi K, Niotis K, Ariza IE, Malviya N, Lee P, Melendez J, Sadek G, Hackett K, Rahman A, Meléndez-Cabrero J, Greer CE, Mosconi L, Krikorian R, Isaacson RS. Sex-Driven Differences in the Effectiveness of Individualized Clinical Management of Alzheimer’s Disease Risk. J Prev Alzheimers Dis. 2022;9(4):731–742. doi: 10.14283/jpad.2022.44. PMID: 36281678.

Sakurai T, Sugimoto T, Akatsu H, Doi T, Fujiwara Y, Hirakawa A, Kinoshita F, Kuzuya M, Lee S, Matsumoto N, Matsuo K, Michikawa M, Nakamura A, Ogawa S, Otsuka R, Sato K, Shimada H, Suzuki H, Suzuki H, Takechi H, Takeda S, Uchida K, Umegaki H, Wakayama S, Arai H; J-MINT study group. Japan-Multimodal Intervention Trial for the Prevention of Dementia: A randomized controlled trial. Alzheimers Dement. 2024 Jun;20(6):3918–3930. doi: 10.1002/alz.13838. Epub 2024 Apr 22. PMID: 38646854; PMCID: PMC11180858.

Salzman T, Sarquis-Adamson Y, Son S, Montero-Odasso M, Fraser S. Associations of Multidomain Interventions With Improvements in Cognition in Mild Cognitive Impairment: A Systematic Review and Meta-analysis. JAMA Netw Open. 2022 May 2;5(5):e226744. doi: 10.1001/jamanetworkopen.2022.6744. PMID: 35503222; PMCID: PMC9066287.

Sandison H, Callan NGL, Rao RV, Phipps J, Bradley R. Observed Improvement in Cognition During a Personalized Lifestyle Intervention in People with Cognitive Decline. J Alzheimers Dis. 2023 Jun 19. doi: 10.3233/JAD-230004. Epub ahead of print. PMID: 37355891.

Sikkes SAM, Tang Y, Jutten RJ, Wesselman LMP, Turkstra LS, Brodaty H, Clare L, Cassidy-Eagle E, Cox KL, Chételat G, Dautricourt S, Dhana K, Dodge H, Dröes RM, Hampstead BM, Holland T, Lampit A, Laver K, Lutz A, Lautenschlager NT, McCurry SM, Meiland FJM, Morris MC, Mueller KD, Peters R, Ridel G, Spector A, van der Steen JT, Tamplin J, Thompson Z; ISTAART Non-pharmacological Interventions Professional Interest Area; Bahar-Fuchs A. Toward a theory-based specification of non-pharmacological treatments in aging and dementia: Focused reviews and methodological recommendations. Alzheimers Dement. 2021 Feb;17(2):255–270. doi: 10.1002/alz.12188. Epub 2020 Nov 20. PMID: 33215876; PMCID: PMC7970750.

Smith GD, Susser E. Zena Stein, Mervyn Susser and epidemiology: observation, causation and action. Int J Epidemiol. 2002 Feb;31(1):34–7. doi: 10.1093/ije/31.1.34. PMID: 11914289.

Soldevila-Domenech N, Ayala-Garcia A, Barbera M, Lehtisalo J, Forcano L, Diaz-Ponce A, Zwan M, van der Flier WM, Ngandu T, Kivipelto M, Solomon A, de la Torre R. Adherence and intensity in multimodal lifestyle-based interventions for cognitive decline prevention: state-of-the-art and future directions. Alzheimers Res Ther. 2025 Mar 17;17(1):61. doi: 10.1186/s13195-025-01691-0. PMID: 40098201; PMCID: PMC11912746.

Soldozy S, Galindo J, Snyder H, Ali Y, Norat P, Yağmurlu K, Sokolowski JD, Sharifi K, Tvrdik P, Park MS, Kalani MYS. Clinical utility of arterial spin labeling imaging in disorders of the nervous system. Neurosurg Focus. 2019 Dec 1;47(6):E5. doi: 10.3171/2019.9.FOCUS19567. PMID: 31786550.

Solomon A, Stephen R, Altomare D, Carrera E, Frisoni GB, Kulmala J, Molinuevo JL, Nilsson P, Ngandu T, Ribaldi F, Vellas B, Scheltens P, Kivipelto M; European Task Force for Brain Health Services. Multidomain interventions: state-of-the-art and future directions for protocols to implement precision dementia risk reduction. A user manual for Brain Health Services-part 4 of 6. Alzheimers Res Ther. 2021 Oct 11;13(1):171. doi: 10.1186/s13195-021-00875-8. PMID: 34635167; PMCID: PMC8507202.

Taherian Fard A, Ragan MA. Modeling the Attractor Landscape of Disease Progression: a Network-Based Approach. Front Genet. 2017 Apr 18;8:48. doi: 10.3389/fgene.2017.00048. PMID: 28458684; PMCID: PMC5394169.

Tessier AJ, Wang F, Korat AA, Eliassen AH, Chavarro J, Grodstein F, Li J, Liang L, Willett WC, Sun Q, Stampfer MJ, Hu FB, Guasch-Ferré M. Optimal dietary patterns for healthy aging. Nat Med. 2025 Mar 24. doi: 10.1038/s41591-025-03570-5. Epub ahead of print. PMID: 40128348.

Tijms BM, Vromen EM, Mjaavatten O, Holstege H, Reus LM, van der Lee S, Wesenhagen KEJ, Lorenzini L, Vermunt L, Venkatraghavan V, Tesi N, Tomassen J, den Braber A, Goossens J, Vanmechelen E, Barkhof F, Pijnenburg YAL, van der Flier WM, Teunissen CE, Berven FS, Visser PJ. Cerebrospinal fluid proteomics in patients with Alzheimer’s disease reveals five molecular subtypes with distinct genetic risk profiles. Nat Aging. 2024 Jan;4(1):33–47. doi: 10.1038/s43587-023-00550-7. Epub 2024 Jan 9. PMID: 38195725; PMCID: PMC10798889.

Toups K, Hathaway A, Gordon D, Chung H, Raji C, Boyd A, Hill BD, Hausman-Cohen S, Attarha M, Chwa WJ, Jarrett M, Bredesen DE. Precision Medicine Approach to Alzheimer’s Disease: Successful Pilot Project. J Alzheimers Dis. 2022;88(4):1411–1421. doi: 10.3233/JAD-215707. PMID: 35811518; PMCID: PMC9484109.

Tricco AC, Lillie E, Zarin W, O’Brien KK, Colquhoun H, Levac D, Moher D, Peters MDJ, Horsley T, Weeks L, Hempel S, Akl EA, Chang C, McGowan J, Stewart L, Hartling L, Aldcroft A, Wilson MG, Garritty C, Lewin S, Godfrey CM, Macdonald MT, Langlois EV, Soares-Weiser K, Moriarty J, Clifford T, Tunçalp Ö, Straus SE. PRISMA Extension for Scoping Reviews (PRISMA-ScR): Checklist and Explanation. Ann Intern Med. 2018 Oct 2;169(7):467–473. doi: 10.7326/M18-0850. Epub 2018 Sep 4. PMID: 30178033.

Tzeng RC, Yang YW, Hsu KC, Chang HT, Chiu PY. Sum of boxes of the clinical dementia rating scale highly predicts conversion or reversion in predementia stages. Front Aging Neurosci. 2022 Sep 23;14:1021792. doi: 10.3389/fnagi.2022.1021792. PMID: 36212036; PMCID: PMC9537043.

Uleman JF, Melis RJF, Hoekstra AG, Olde Rikkert MGM, Quax R; Australian Imaging, Biomarker and Lifestyle study of Aging and Alzheimer’s Disease Neuroimaging Initiative studies. Exploring the potential impact of multi-factor precision interventions in Alzheimer’s disease with system dynamics. J Biomed Inform. 2023 Sep;145:104462. doi: 10.1016/j.jbi.2023.104462. Epub 2023 Jul 27. PMID: 37516375.

Uleman JF, Quax R, Melis RJF, Hoekstra AG, Olde Rikkert MGM. The need for systems thinking to advance Alzheimer’s disease research. Psychiatry Res. 2024 Mar;333:115741. doi: 10.1016/j.psychres.2024.115741. Epub 2024 Jan 17. PMID: 38277813.

Wainberg M, Magis AT, Earls JC, Lovejoy JC, Sinnott-Armstrong N, Omenn GS, Hood L, Price ND. Multiomic blood correlates of genetic risk identify presymptomatic disease alterations. Proc Natl Acad Sci U S A. 2020 Sep 1;117(35):21813–21820. doi: 10.1073/pnas.2001429117. Epub 2020 Aug 19. PMID: 32817414; PMCID: PMC7474629.

Yaffe K, Vittinghoff E, Dublin S, Peltz CB, Fleckenstein LE, Rosenberg DE, Barnes DE, Balderson BH, Larson EB. Effect of personalized risk-reduction strategies on cognition and dementia risk profile among older adults: the SMARRT randomized clinical trial. JAMA Intern Med. 2023:e236279. 10.1001/jamainternmed.2023.6279. Epub ahead of print. PMID: 38010725; PMCID: PMC10682943.

Yang J, Dong Y, Yan S, Yi L, Qiu J. Which Specific Exercise Models Are Most Effective on Global Cognition in Patients with Cognitive Impairment? A Network Meta-Analysis. Int J Environ Res Public Health. 2023 Feb 4;20(4):2790. doi: 10.3390/ijerph20042790. PMID: 36833483; PMCID: PMC9957167.

Yang QH, Lyu X, Lin QR, Wang ZW, Tang L, Zhao Y, Lyu QY. Effects of a multicomponent intervention to slow mild cognitive impairment progression: A randomized controlled trial. Int J Nurs Stud. 2022 Jan;125:104110. doi: 10.1016/j.ijnurstu.2021.104110. Epub 2021 Oct 10. PMID: 34736073.

Zhang Y, Chen H, Li R, Sterling K, Song W. Amyloid β-based therapy for Alzheimer’s disease: challenges, successes and future. Signal Transduct Target Ther. 2023 Jun 30;8(1):248. doi: 10.1038/s41392-023-01484-7. PMID: 37386015; PMCID: PMC10310781.

Zhang X, Fu Z, Meng L, He M, Zhang Z. The Early Events That Initiate β-Amyloid Aggregation in Alzheimer’s Disease. Front Aging Neurosci. 2018 Nov 13;10:359. doi: 10.3389/fnagi.2018.00359. PMID: 30542277; PMCID: PMC6277872.

Zülke AE, Pabst A, Luppa M, Roehr S, Seidling H, Oey A, Cardona MI, Blotenberg I, Bauer A, Weise S, Zöllinger I, Sanftenberg L, Brettschneider C, Döhring J, Lunden L, Czock D, Haefeli WE, Wiese B, Hoffmann W, Frese T, Gensichen J, König HH, Kaduszkiewicz H, Thyrian JR, Riedel-Heller SG. A multidomain intervention against cognitive decline in an at-risk-population in Germany: Results from the cluster-randomized AgeWell.de trial. Alzheimers Dement. 2024 Jan;20(1):615–628. doi: 10.1002/alz.13486. Epub 2023 Sep 28. PMID: 37768074; PMCID: PMC10917033.

Zülke AE, Riedel-Heller SG, Wittmann F, Pabst A, Röhr S, Luppa M. Gender-Specific Design and Effectiveness of Non-Pharmacological Interventions against Cognitive Decline - Systematic Review and Meta-Analysis of Randomized Controlled Trials. J Prev Alzheimers Dis. 2023;10(1):69–82. doi: 10.14283/jpad.2022.80. PMID: 36641611.

