## Supplemental Appendix 1 for "Multidomain Therapy for Alzheimer’s Disease: A Scoping Review"

### ASSESSMENTS OF TRIAL INFORMATION FOR TABLES 1 & 2

We assessed available information for each trial to best provide summary values for Table 1. This information is mostly derived from publications (e.g., published protocols and results) and information provided in clinicaltrials.gov. We used FAST as a unifying variable for dementia stage, as it is more precise (i.e., has more tiers) than other options and staging can be approximated based on a variety of assessments. Such unification is needed; few trials use the same assessments for their inclusion/exclusion criteria. Trials with inclusion criteria that included all older individuals without any other AD-relevant criteria were excluded (see also **Supplemental Appendix 2**) as we considered these to not be “Alzheimer’s disease” (AD) trials. Trials that included AD risk factors (other than age) as inclusion criteria were included even if most or all participants were FAST stage 1; we considered these to be trials of presymptomatic AD.

Estimates of relative effect size and Cohen’s d effect size were often difficult; authors rarely report effect sizes. Cohen’s d (or nearly equivalently Hedge’s g) effect sizes were available from publications for COMBAT & FINGER; we used these three reported values in Table 2. For all other cases we did our best to estimate Cohen’s d based on the means of the two arms and the standard deviation of the outcome measure. Means of outcome measures were almost universally available from publications; standard deviations in many cases had to be estimated. As a result, our estimates for Cohen’s d might be off by a factor of 2 or more. However, such imprecision has little effect on any perspective or conclusion we present. Our estimates of relative effect size also suffer from imprecision for two reasons: (1) the precise meaning and interpretation of “relative effect size" varies depending on the type of variable (e.g., normalized or linearized) and the directional change from baseline of the two trial arms, and (2) absolute values of the primary outcome variables are not reported in all publications. Generally speaking, relative effect sizes are deprecated as statistics. However, they are often reported in the context of AD clinical trials and a subset of readers will expect to see them. One of our goals for computing and tabulating relative effect sizes in Table 2 is to demonstrate their unreliability for most purposes, including comparative purposes.

**AgeWell.de.** Participants in AgeWell.de had a mean Montreal Cognitive Assessment (MoCA) score of 24.5 points at baseline, consistent with a median FAST score of 1.

**Aliabad-e Katul.** Elderly women (ages 60 to 85 years) with MCI in Iran (Damirchi et al., 2018) were recruited as assessed by MMSE 18-23 (approximately FAST stage 4-5).

**Bae (aka, Effect of the population-based approach for prevention of dementia).** We refer to this trial (dementia (UMIN-CTR Clinical Trial UMIN000026479) with the shorthand "Bae", the first author of the 2019 results paper (Bae et al., 2019), consistent with the nomenclature of Soldevila-Domenech et al. (2025). Inclusion criteria included diagnosis with mild cognitive impairment, consistent with FAST stage 3.

**BioRAND.** BioRAND (Niotis et al., 2025; https://ind.org/newsroom/the-biorepository-study-for-neurodegenerative-diseases) is a cohort with ongoing recruitment. Metadata has yet to be published in a full peer reviewed manuscript. Inclusion criteria include subjects with family history of NDD with no/minimal neurological symptoms, which we represent as FAST stage 1. Numbers of participants are expected to increase over time; duration of participation is expected to increase and vary between participants. Individuals are non-randomly designated as cases and controls.

**CEDAR.** CEDAR aimed to recruit participants at risk for AD dementia. Inclusion criteria included: (1) Family history of Alzheimer's disease (AD), and (2) No or minimal cognitive complaints. Exclusion criteria included MCI or dementia. Some individuals with SCI may have been FAST stage 2. We summarize this in Table 1 as FAST Stage 1 at enrollment.

**COCOA.** At baseline, 26% of COCOA participants were FAST 2, 40% were FAST 3, and 33% were FAST 4 (Roach et al., 2023). We summarize this in Table 1 as FAST Stage 2 at enrollment.

**COMBAT.** Inclusion criteria included having subjective cognitive complaints as indicated by the self-rated Ascertain Dementia 8-Item Questionnaire (AD-8) score greater than or equal to 2 (Liu et al., 2025), consistent with FAST stage 2. Participants in the modified intention to treat cohort had an average age of 69.7 years in the intervention arm and 73.1 years in the control arm.

**ENLIGHTEN.** ENLIGHTEN aimed to recruit older adults with cognitive impairment not dementia (CIND) who were at risk for further cognitive decline due to their sedentary lifestyle and cardiovascular risk factors. Inclusion criteria included: (1) subjective cognitive complaints: Score of ≥0.5 on the Mail-in Cognitive Function Screening Instrument, and (2) objective cognitive impairment: Either a score of 19-25 on the Montreal Cognitive Assessment (MoCA) or a score of ≤12 on letter fluency or ≤15 on animal fluency. Exclusion criteria included dementia. We summarize this in Table 1 as FAST Stage 2-3 at enrollment.

**Evanthea.** Inclusion criteria for the Evanthea trial (clinicaltrials.gov/study/NCT05894954) included cognitive impairment or early-stage dementia as demonstrated by combination of AQ-21 score >4 and either MoCA 18-26 or greater than or equal 2 scores in the bottom 50th percentile for NCI or Executive Function, Verbal, Visual, or Composite sub-tests. We summarize this in Table 1 as FAST Stage 3 at enrollment.

**FINGER.** FINGER aimed to recruit participants from previous population-based survey cohorts in Finland, specifically targeting individuals at increased risk of cognitive decline but without substantial cognitive impairment. These cognitive criteria selected individuals with cognitive performance at the mean level or slightly lower than expected for their age according to Finnish population norms, but without substantial cognitive decline (Ngandu et al., 2015). Participants had to meet at least one of the following criteria at the screening visit: (1) Word List Learning task (10 words × 3) ≤19 words, (2), Word List Recall ≤75%, (3) MMSE ≤26/30 points. A participant at the mean cognitive performance for their age would be FAST 1, but some participants could have had subjective impairment (FAST 2), and some scores on the MMSE or other tests could have evidenced objective impairment (FAST 3). We summarize this in Table 1 as FAST Stage 1-2 at enrollment.

**ITHNCLR.** Inclusion criteria for the ITHNCLR study included a MoCA score of 12–23. We summarize this in Table 1 as FAST Stage 3-4 at enrollment.

**LETHE.** Inclusion criteria for the LETHE trial were increased dementia risk and no substantial cognitive impairment. We summarize this in Table 1 as FAST Stage 1 at enrollment.

**LIEAD.** Inclusion criteria for the Lifestyle Intervention for Early Alzheimer's Disease (LIEAD) (Ornish-D et al., 2024) trial included a diagnosis of MCI or early dementia due to AD process, with a MoCA score of 18 or higher (National Institute on Aging–Alzheimer's Association McKhann and Albert 2011 criteria); moderate of severe dementia was excluded. We summarize this in Table 1 as FAST Stage 5 at enrollment.

**MAPT.** The French Multidomain Alzheimer Preventive Trial (MAPT) aimed to enroll frail elderly individuals with good functional and cognitive status but at risk of cognitive decline.

Participants were recruited through advertisements, general practitioners, and memory clinics in France. Inclusion criteria included at least one of: (1) Spontaneous memory complaint expressed to a general practitioner, (2) Limitation in one instrumental activity of daily living (IADL), (3) slow walking speed. Exclusion criteria included: (1) MMSE score < 24, and (2) dementia. Some of these individuals with just slow walking gait could be FAST 1, and some with low MMSE scores might be FAST 3. We summarize this in Table 1 as FAST Stage 2 at enrollment.

**MYB.** The Maintain Your Brain (MYB) trial was an internet-based study involving over 6,000 participants from the Sax Institute's 45 and Up Study in Australia. Inclusion criteria for MYB included dementia risk factors (e.g., high blood pressure or less than 13 years of education); exclusion criteria included dementia. We summarize this in Table 1 as FAST Stage 1 at enrollment.

**PREVENTION.** At baseline, 22% of PREVENTION participants were FAST 2, 77% were FAST 3, and 21% were FAST 4. We summarize this in Table 1 as FAST Stage 3 at enrollment.

**RECODE.** Inclusion criteria for the Reversal of Cognitive Decline (RECODE) study (Toups et al., 2022) included (1) cognitive impairment, as demonstrated by a combination of Alzheimer's Questionnaire (AQ21)>5 and either Montreal Cognitive Assessment (MoCA) of 19–26 or CNS Vital Signs <50th percentile in at least two subtests or <70th percentile for the Neurocognitive Index (NCI). Exclusion criteria included MoCA score <19. We summarize this in Table 1 as FAST Stage 3 at enrollment.

**SMARRT.** Inclusion criteria for SMARRT (Yaffe et al., 2023) included low-to-normal cognitive performance based on Brief CASI score 25 to 32, inclusive. We summarize this in Table 1 as FAST Stage 1 at enrollment. Inclusion criteria also included high risk for dementia based on ≥2 of 8 targeted risk factors: (1) physical inactivity, (2) uncontrolled hypertension, (3) poor sleep, (4) taking a prescription medication that may adversely affect cognition, (5) high depressive symptoms, (6) uncontrolled diabetes, (7) social isolation, and (8) currently smoking.

**The Taiwan Health promotion Intervention Study for Community Elders Efficacy Study (THISCE-EFF).** THISCE-EFF was a 2014 Taiwanese 2014 study (Chen et al., 2020). Inclusion criteria included subjective memory impairment (SCI) and/or loss of ≥ 1 instrumental activities of daily living (IADL), and/or timed 6 m walk speed ≤ 1 m/s. It is not clear from the Chen et al. report how many enrolled individuals met the option of SCI, but it seems reasonable to estimate that this cohort had an average FAST stage 1.

**US POINTER.** Inclusion criteria for the Alzheimer's Association U.S. Study to Protect Brain Health Through Lifestyle Intervention to Reduce Risk (U.S. POINTER) included a Modified Telephone Interview for Cognitive Status (mTICS) score >32 and Global Clinical Dementia Rating (CDR) scale score ≤0.5. We summarize this in Table 1 as FAST Stage 1 at enrollment.

**Xinzhuang.** Inclusion criteria for the trial reported by Fan et al. (2024), which took place in Xinzhuang, included diagnosis of MCI according to the Peterson criteria: (1) Cognitive concern or complaint by the subject or proxy, with Clinical Dementia Rating (CDR) = 0.5, (2) Objective impairment in at least 1 cognitive domain (performance 1.5 SD below the mean using Chinese population norms), (3) normal functional activities, and (4) Absence of dementia. We summarize this in Table 1 as FAST Stage 3 at enrollment.

**Yang (aka, “A study examining the effectiveness of a multi-modal intervention to slow down the progression of mild cognitive impairment”).** We refer to this trial (Chinese clinical trial registration number ChiCTR1900026042) with the shorthand “Yang”, the first author of the

2022 results paper (Yang et al., 2022), consistent with the nomenclature of Soldevila-Domenech et al. (2025). Inclusion criteria included diagnosis with mild cognitive impairment, consistent with FAST stage 3.

**Additional References not Cited in Main Text**

Ngandu T, Lehtisalo J, Solomon A, Levälahti E, Ahtiluoto S, Antikainen R, Bäckman L, Hänninen T, Jula A, Laatikainen T, Lindström J, Mangialasche F, Paajanen T, Pajala S, Peltonen M, Rauramaa R, Stigsdotter-Neely A, Strandberg T, Tuomilehto J, Soininen H, Kivipelto M. A 2 year multidomain intervention of diet, exercise, cognitive training, and vascular risk monitoring versus control to prevent cognitive decline in at-risk elderly people (FINGER): a randomised controlled trial. Lancet. 2015 Jun 6;385(9984):2255-63. doi: 10.1016/S0140-6736(15)60461-5. Epub 2015 Mar 12. PMID: 25771249.
