## Supplemental Appendix 2 for "Multidomain Therapy for Alzheimer’s Disease: A Scoping Review"

### INCLUSION AND EXCLUSION OF STUDIES

**PubMed Boolean Search String:**

```
(((("subjective cognitive decline" OR SCD OR "subjective memory impairment" OR "subjective cognitive impairment" OR SCI OR prefrail OR "Cognitive Dysfunction/prevention and control"[MAJR])) OR (("mild cognitive impairment" OR MCI OR (prodromal AND ("alzheimer disease"[MeSH Terms])))) AND (((alzheimer AND disease) OR "alzheimer disease" OR "Alzheimer's disease" OR "mild cognitive impairment" OR prefrail OR "Cognitive Dysfunction/prevention and control"[MAJR])) )) AND (multidomain OR multimodal OR "multi-domain" OR "multi-modal" OR "multi-component" OR multicomponent OR multidimensional OR multisystem OR "combined training" OR "combined modality therapy") AND (trial OR (randomized AND study)) AND (diet OR dietary OR exercise OR physical OR "combined modality therapy")
```

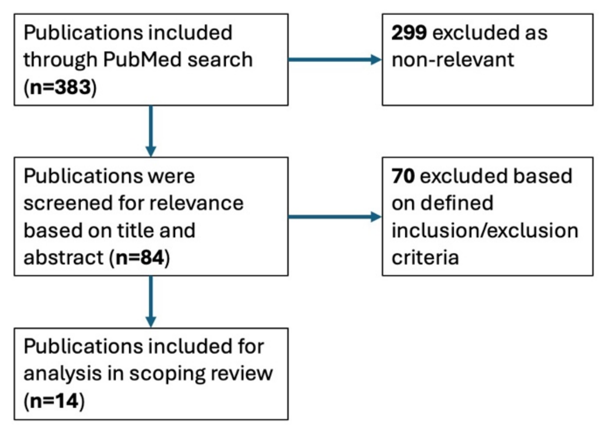


**Scoping Review Screening Flowchart.**

Many of the studies screened out of our scoping analysis are nevertheless informative with respect to multidomain interventions for ADRD. Many of them focus on a single intervention such as diet or exercise that affects multiple domains. Likewise, we did not include trials that only employed pharmaceuticals, even if multiple pharmaceuticals were used. In principle, a multi-pharmaceutical trial could be multidomain; we expect to see such trials in the future. We do not include the "combination therapy" of concomitant use of memantine and cholinesterase inhibitors (ChEIs), as it has been adequately reviewed elsewhere, and the largely/entirely symptomatic mechanisms of actions of these drugs are well understood (Kabir et al., 2020). We do not include combinations of anti-tau and anti-amyloid drugs in this review, as this combination is new and arguably both drugs target the same domain; we expect more reviews on such combinations to be published in the future (Cummings et al., 2024). We also note that from a purely pedantic point of view, any single intervention will always target multiple domains; it is impossible for any intervention not to affect a myriad of biopsychosocial subsystems, at least in imperceptibly small ways; we restrict our discussion to those effects that are important, recognizing that our current assessment of importance is nascent. We did not include observational studies without interventions (i.e., "registries"), although we did include observational studies of interventions without control arms (but not in **Table 2**). We briefly describe a subset of trials we did not include below.

Our primary focus, for purposes of this review, was multidomain interventions. Therefore, for studies with more than two arms (e.g., MAPT, Aliabad-e Katul, Deutschschweiz, SFIT), we ignored unimodal arms, consistent with our exclusion of unimodal trials in general. We do not summarize here overall ANOVA significance for multiple-arm statistics reported by those authors. We report only the pairwise statistics between the multidomain arm(s) and the control arm for these studies. This choice may be important, as it results in our reporting in our **Table 2** of the MAPT (multidomain portion) arm 1 trial result as significant: p=0.047 as reported in Table 2 of Andrieu et al. (2017), despite the statement in the Abstract of Andrieu et al.: "The multidomain intervention and polyunsaturated fatty acids, either alone or in combination, had no significant effects on cognitive decline over 3 years in elderly people with memory complaints."

The Boolean search term for the concept of "Alzheimer's disease" was necessarily broad. Many people prefer not to be labeled with the term "Alzheimer's". This can be particularly true in presymptomatic, early, or prodromal disease. Furthermore, some practitioners at the time of their study design may have considered a connotation of the term “Alzheimer’s disease” to be solely restricted to advanced stages of the disease with loss of function, and not to early stages, which they may term SCI or MCI (for example). To maximize recruitment to an early-stage AD trial, it may be best not to use the term “Alzheimer’s” as part of the trial name or description. This, among other reasons, motives our broad inclusion of terms (e.g., ‘prefrail’) in an initial Boolean search, with subsequent screening of the participant inclusion/exclusion criteria for each trial to determine if that trial meets the scoping review manuscript inclusion/exclusion criteria.

### EXLCUDED STUDIES

We summarize a select set of excluded studies, focusing on those that came close to meeting inclusion criteria.

**A Multi-domain Lifestyle Intervention Among Aged Community-residents in Zhejiang, China (HERITAGE).** HERITAGE is a 2-trial that explores the efficacy of a multidomain intervention among 1200 elderly residents with a higher risk of cognitive decline in China. Results are pending (NCT05256121).

**A Multidomain Intervention Program for Older People with Dementia (Hanoi**). This trial based in Vietnam to assess the feasibility of a multidomain intervention for older people with dementia in nursing homes is registered with ClinicalTrials.gov (NCT04948450).

**A Study on the Effectiveness of Multidomain Intervention Program for Reducing Risks of Dementia (Seoul).** Results from this multimodal study are pending (NCT03786510).

**Aliabad-e Katul.** Damirchi et al. (2018) conducted an 8-week 4-arm RCT at Aliabad-e Katul, Golestan Province, Iran to tests to the effects of cognitive training, exercise, or both (N=13), compared to a control arm (N=9). In elderly women (ages 60 to 85 years) with MCI defined by MMSE 18-23 (approximately FAST 4-5) and assessed several outcome measures including the Wechsler Adult Intelligence Scale-Third Edition (WAIS III). Average baseline assessment values for the control arm were lower than baseline assessments for the other arms. Results were not adjusted for baseline values. The authors do not report a significant difference between the multidomain arm and the control arm. We include this study in **Table 1**, but due to the difficulty in computing effect size and significance for the difference between the multidomain and control arms, we do not include it in **Table 2**.

**ASPIS.** ASPIS was an Austrian RCT focused on post-stroke cognitive decline (Brainin et al., 2015).

**Complex Mental and Physical Activity in Older Women.** The Complex Mental and Physical Activity in Older Women and Cognitive Performance trial (Klusmann et al., 2010) was a 6-month RCT in Germany to compare the effects of exercise to the effects of cognitive training on cognitive performnce. It compared these as unimodal interventions to each other and to a control arm. Both groups showed improvement with respect to controls.

**Dementia - Early Detection and Intervention (DEDI).** The DEDI trial (ClinicalTrials.gov NCT04440969) was a 24-week multidomain RCT (Ng et al., 2021) with 199 participants >54 years in 2018. Participants were excluded if they were those diagnosed with cognitive disorders. In other contexts, this trial has been referred to with the shorthand "Ng", the first author of the 2024 results paper, consistent with the nomenclature of Soldevila-Domenech et al. (2025).

**Deutschschweiz.** This 5-week 3-arm RCT (Brasser et al., 2022) enrolled participants into a multidomain intervention (N=41), a cognitive training intervention, and a control arm (N=41). All older adults in German-speaking Switzerland (aka, Deutschschweiz) ages 65-90 years with access to a computer, email, and a printer were eligible. The majority participants are unlikely to have AD, except due to background incidence of AD in this population. Participants were recruited in 2021. The interventions (physical, cognitive, social, mindful, coordination, and creative activation) were remote, as this was during to the COVID pandemic. Multiple outcomes were measured, including the RAVLT. At 5-weeks, the multidomain arm performance was significantly better than the control ($p < 0.01$) at working memory.

**Dutch Multidomain Lifestyle Intervention in Older Adults at Risk of Cognitive Decline (FINGER-NL).** FINGER-NL is a multi-center, randomized, controlled, multidomain lifestyle intervention trial among 1,206 older adults at risk for cognitive decline with a duration of 24 months (Deckers et al., 2024). Results are pending (NCT05256199).

**Effectiveness of a Lifestyle Multidomain Intervention to Prevent Cognitive Decline in the Basque Country (GOIZ ZAINDU).** GOIZ ZAINDU was a pilot study to adapt the FINGER methodology to the Basque population (Tainta et al., 2024). Primary outcomes were retention rate and adherence to the intervention program. Secondary outcomes showed cognitive benefit in the multidomain intervention group (NCT04840030 & NCT06163716).

**eMIND.** eMIND was a 2017 6-month pilot study to test the feasibility and compliance to a web-based multidomain intervention in France in 120 participants aged 65+ years with FAST stage 2 at baseline. Cognitive outcome measures were included only as secondary measures. Differences in these measures were not reported as significant (de Souto et al., 2021).

**HATICE.** HATICE was a 2015 18-month multidomain RCT. The 2724 participants were older adults at risk for cardiovascular disease with FAST stage 1. The primary outcome measure was a composite score of systolic blood pressure, LDL cholesterol, and body-mass index (BMI). Secondary outcomes included individual risk factors and cardiovascular endpoints (Richard et al., 2019). MMSE and a composite cognitive measure were also reported. Differences in these measures were not reported as significant.

**Human-APPlication Combined Approach for Prevention of Alzheimer's Disease (HAPPCAP-AD).** HAPPCAP-AD (Ganmore et al., 2020) tests a personalized 18-months intervention for prevention of cognitive decline in middle-aged individuals at high AD risk due to a parental family history. Multimodal intervention is provided via a smartphone application (app) with study team guidance through phone-calls. Results are pending (NCT05256121).

**J-MIND-Diabetes.** J-MIND-Diabetes was a Japanese RCT to study the effect of a multidomain intervention in older adults with diabetes and cognitive decline (Sugimoto et al., 2024).

**MAPT.** MAPT was a 2008 RCT in France with 1680 participants. Two of the arms included omega-3 polyunsaturated fatty acids (800 mg docosahexaenoic acid and 225 mg eicosapentaenoic acid daily) coupled to the hypothesis that this omega-3 unimodal intervention had an independent causal effect. The overall purpose of the MAPT trial was designed in large part to test this specific unimodal supplement intervention. Therefore it is problematic to incorporate results from the MAPT trial into a review of prospective RCTs with pre-declared hypotheses that multidomain interventions improve AD. One conceivable retrospective approach to these data would be to combine the data from the two arms that included multidomain interventions and compare these data to the placebo arm. However, such an analysis is not reported in Andrieu et al. (2019). Raw p-values reported by Andrieu et al. for comparison of these two arms individually to placebo were reported as 0.047 and 0.090. The authors and some subsequent review articles have summarized these results as, "The multidomain intervention and polyunsaturated fatty acids, either alone or in combination, had no significant effects on cognitive decline over 3 years in elderly people with memory complaints." We believe this is too strong a statement, at least without further context. We therefore include results from MAPT in Table 1 and Table 2 as a separate row for each multidomain arm, with the addition of a footnote.

**Meng (aka, Construction and effectiveness of multidomain dementia prevention program based on life-course model of modifiable risk factors to dementia).** We refer to this trial (Chinese clinical trial registration number ChiCTR2100053417) with the shorthand "Meng", the first author of the 2024 results paper (Meng et al., 2024), consistent with the nomenclature of Soldevila-Domenech et al. (2025). This RCT focused on multidomain interventions for healthy older adults.

**MIND-AD$_{\text{mini}}$.** MIND-AD mini was a 2017 6-month European RCT (Thunborg et al., 2024). The 93 participants were 60–85 years old and had early AD (baseline FAST ~3). Primary outcomes focused on retention. Cognitive outcome measures including the CDR-SB were included only as secondary measures. The relative risk of cognitive decline assessed by the CDR-SB was reported and was significant for only one of two multimodal arms (p =~ 0.025), showing a relative improvement in the intervention arm of ~25% (effect size was difficult to estimate from reported data).

**Park.** We refer to this trial with the shorthand "Park", the first author of the 2019 results paper (Park et al., 2019), consistent with the nomenclature of Soldevila-Domenech et al. (2025). This RCT focused on multidomain interventions for healthy older adults (Mini-Mental State Examination (MMSE) of >–1.5 SD).

**PASSWORD Study – Promoting Safe Walking.** The primary endpoint of PASSWORD (www.isrctn.com/ISRCTN52388040) was gait speed. Cognitive training and exercise were used in this multidomain study, and cognitive outcomes such as STROOP were measured. Healthy older adults were recruited (not specifically AD). Benefit on cognitive outcomes was observed from a synergistic combination of interventions (Sipilä et al., 2021).

**PreDIVA.** Prevention of Dementia by Intensive Vascular Care (PreDIVA) was large (3526 participants), multidomain lifestyle prevention trial conducted in the Netherlands (Moll van Charante et al., 2016). It focused on dementia outcomes but without explicit mention of AD.

Primary outcomes were cumulative incidence of dementia and disability score (Academic Medical Center Linear Disability Score [ALDS]) at 6 years of follow-up. The investigators use the term 'multidomain' to describe a multimodal largely cardiovascular intervention, but that also targeted metabolic domains: participants were randomized to either a multimodal intervention that targeted vascular risk factors (smoking, unhealthy diet, physical inactivity, overweight, hypertension, dyslipidemia, and diabetes) by a practice nurse or to usual care. According to the investigators, the PreDIVA intervention did not result in a reduced incidence of all-cause dementia. The investigators speculate that their study was underpowered due to low baseline cardiovascular risks and high standards of usual care—both with the effect of creating a control group not likely to diverge from an intervention arm, even if the intervention was effective.

**Reducing Dementia Risk with Digital Health Coaching (DC-MARVEL).** DC-MARVEL is a 2-year randomized controlled trial on dementia prevention. The purpose of this study is to determine the effect of a digital cognitive health program on dementia risk, cognitive function, and general health outcomes in middle age to older adults compared to a control group that receives health education. Results are pending (NCT04559789).

**Singapore Frailty Intervention Trial (SFIT).** SFIT was a 6-month 5-arm RCT in Singapore in 2009. One of these arms was multidomain (N=49), one was control (N=50) for participants >64 years old. Participants were cognitively normal (Mini-Mental State Examination >23). Primary outcome was frailty (Ng et al., 2015). An exploratory analysis of cognitive function was also reported (Ng et al., 2018).

**SINgapore GERiatric Intervention Study to Reduce Cognitive Decline and Physical Frailty (SINGER).** The SINGER Trial is a FINGER-inspired trial based in Singapore (Xu et al., 2022). Results are pending (NCT05007353).

**SINGER-pilot.** SINGER-pilot (Chew et al., 2021) was a 6-month 2018 pilot study for the SINGER Trial . It was a 2-arm RCT with 70 participants >64 years old. Both arms were multidomain (no control arm). Inclusion criteria included mild-to-moderate frailty as defined by a Physical Performance Test (PPT) score of 17 to 32. Primary outcome measures were related to logistics and feasibility.

**StayFitLonger.** StayFitLonger was a 26 week multinational RCT that tested a multidomain intervention on cognitively normal (≥ 26 on the Montreal Cognitive Assessment) older adults (Belleville et al., 2023). Participants receiving the multidomain intervention showed improvement in global cognition.

**SUPERBRAIN.** SUPERBRAIN (Moon et al., 2021) was 3-arm RCT to test either a facility- or home-based multimodal intervention in 152 participants who were cognitively normal at baseline (MMSE Z score of ≥ -1.5). Participants were 60-79 years of age. The intervention lasted 24 weeks. It comprised vascular risk management, cognitive training, social activity, physical exercise, nutrition guidance, and motivational enhancement.

**SYNchronizing, Exercises and Remedies to GaIn Cognition@home (SYNERGIC-2).** The SYNERGIC-2 Trial is an actively recruiting member of the WW-FINGERS consortium based in Canada (NCT05375513).

**The Taiwan Health promotion Intervention Study for Community Elders Empowerment Study (THISCE-EMP).** THISCE-EFF and THISCE-EMP were two separate Taiwanese 2014 studies (Chen et al., 2020). They are sometimes described together as “THISCE”. These had non-overlapping participants and the protocols differed. THISCE-EMP did not include a control arm, so we only include THISCE-EFF in our main text tabulations.

**WW-FINGERS.** World-Wide FINGERS (WW-FINGERS) is a global network of studies adapting the FINGER model to diverse populations and contexts worldwide. As this is a consortium, we report on component trials with published data. No WW-FINGERS meta-analysis is yet available.

**Additional References not Cited in Main Text**


Belleville S, Cuesta M, Bieler-Aeschlimann M, Giacomino K, Widmer A, Mittaz Hager AG, Perez-Marcos D, Cardin S, Boller B, Bier N, Aubertin-Leheudre M, Bherer L, Berryman N, Agrigoroaei S, Demonet JF. Pre-frail older adults show improved cognition with StayFitLonger computerized home-based training: a randomized controlled trial. Geroscience. 2023 Apr;45(2):811-822. doi: 10.1007/s11357-022-00674-5. Epub 2022 Oct 21. Erratum in: Geroscience. 2023 Oct;45(5):3099-3100. doi: 10.1007/s11357-023-00868-5. PMID: 36266559; PMCID: PMC9589849.

Brainin M, Matz K, Nemec M, Teuschl Y, Dachenhausen A, Asenbaum-Nan S, Bancher C, Kepplinger B, Oberndorfer S, Pinter M, Schnider P, Tuomilehto J; ASPIS Study Group. Prevention of poststroke cognitive decline: ASPIS--a multicenter, randomized, observer-blind, parallel group clinical trial to evaluate multiple lifestyle interventions--study design and baseline characteristics. Int J Stroke. 2015 Jun;10(4):627-35. doi: 10.1111/ijs.12188. Epub 2013 Nov 10. PMID: 24206541.

Brasser M, Frühholz S, Schneeberger AR, Ruschetti GG, Schaerli R, Häner M, Studer-Luethi B. A Randomized Controlled Trial Study of a Multimodal Intervention vs. Cognitive Training to Foster Cognitive and Affective Health in Older Adults. Front Psychol. 2022 Jun 20;13:866613. doi: 10.3389/fpsyg.2022.866613. PMID: 35795412; PMCID: PMC9251428.

Cummings JL, Osse AML, Kinney JW, Cammann D, Chen J. Alzheimer's Disease: Combination Therapies and Clinical Trials for Combination Therapy Development. CNS Drugs. 2024 Aug;38(8):613-624. doi: 10.1007/s40263-024-01103-1. Epub 2024 Jun 27. PMID: 38937382; PMCID: PMC11258156.

Deckers K, Zwan MD, Soons LM, Waterink L, Beers S, van Houdt S, Stiensma B, Kwant JZ, Wimmers SCPM, Heutz RAM, Claassen JAHR, Oosterman JM, de Heus RAA, van de Rest O,

Vermeiren Y, Voshaar RCO, Smidt N, Broersen LM, Sikkes SAM, Aarts E; MOCIA consortium; FINGER-NL consortium; Köhler S, van der Flier WM. A multidomain lifestyle intervention to maintain optimal cognitive functioning in Dutch older adults-study design and baseline characteristics of the FINGER-NL randomized controlled trial. Alzheimers Res Ther. 2024 Jun 13;16(1):126. doi: 10.1186/s13195-024-01495-8. PMID: 38872204; PMCID: PMC11170777.

de Souto Barreto P, Pothier K, Soriano G, Lussier M, Bherer L, Guyonnet S, Piau A, Ousset PJ, Vellas B. A Web-Based Multidomain Lifestyle Intervention for Older Adults: The eMIND Randomized Controlled Trial. J Prev Alzheimers Dis. 2021;8(2):142-150. doi: 10.14283/jpad.2020.70. PMID: 33569560; PMCID: PMC7754697.

Ganmore, I., Ravona-Springer, R., Livny, A., Matatov, A., Ziat, A., Bem-Moshe, A. and Beeri, M.S. (2020), HAPPCAP-AD (Human-Application Combined Approach for Prevention of Alzheimer's Disease): A novel feasibility study of a personalized midlife intervention to prevent AD. Alzheimer's Dement., 16: e044234. https://doi.org/10.1002/alz.044234

Kabir MT, Uddin MS, Mamun AA, Jeandet P, Aleya L, Mansouri RA, Ashraf GM, Mathew B, Bin-Jumah MN, Abdel-Daim MM. Combination Drug Therapy for the Management of Alzheimer's Disease. Int J Mol Sci. 2020 May 5;21(9):3272. doi: 10.3390/ijms21093272. PMID: 32380758; PMCID: PMC7246721.

Klusmann V, Evers A, Schwarzer R, Schlattmann P, Reischies FM, Heuser I, Dimeo FC. Complex mental and physical activity in older women and cognitive performance: a 6-month randomized controlled trial. J Gerontol A Biol Sci Med Sci. 2010 Jun;65(6):680-8. doi: 10.1093/gerona/glq053. Epub 2010 Apr 23. PMID: 20418350.

Meng X, Su J, Gao T, Ma D, Zhao Y, Fang S, Zhi S, Li H, Sun J. Multidomain interventions based on a life-course model to prevent dementia in at-risk Chinese older adults: A randomized controlled trial. Int J Nurs Stud. 2024 Apr;152:104701. doi: 10.1016/j.ijnurstu.2024.104701. Epub 2024 Jan 26. PMID: 38330865.

Moll van Charante EP, Richard E, Eurelings LS, van Dalen JW, Ligthart SA, van Bussel EF, Hoevenaar-Blom MP, Vermeulen M, van Gool WA. Effectiveness of a 6-year multidomain vascular care intervention to prevent dementia (preDIVA): a cluster-randomised controlled trial.

Lancet. 2016 Aug 20;388(10046):797-805. doi: 10.1016/S0140-6736(16)30950-3. Epub 2016 Jul 26. PMID: 27474376.

Moon SY, Hong CH, Jeong JH, Park YK, Na HR, Song HS, Kim BC, Park KW, Park HK, Choi M, Lee SM, Chun BO, Koh SH, Park SA, Park HH, Jin JH, Lee EH, Kim SM, Han SM, Kim JS, Ha J, Choi SH. Facility-based and home-based multidomain interventions including cognitive training, exercise, diet, vascular risk management, and motivation for older adults: a randomized controlled feasibility trial. Aging (Albany NY). 2021 Jun 18;13(12):15898-15916. doi: 10.18632/aging.203213. Epub 2021 Jun 18. PMID: 34148030; PMCID: PMC8266338.

Ng PEM, Nicholas SO, Wee SL, Yau TY, Chan A, Chng I, Yap LKP, Ng TP. Implementation and effectiveness of a multi-domain program for older adults at risk of cognitive impairment at neighborhood senior centres. Sci Rep. 2021 Feb 15;11(1):3787. doi: 10.1038/s41598-021-83408-5. PMID: 33589714; PMCID: PMC7884402.

Ng TP, Feng L, Nyunt MS, Feng L, Niti M, Tan BY, Chan G, Khoo SA, Chan SM, Yap P, Yap KB. Nutritional, Physical, Cognitive, and Combination Interventions and Frailty Reversal Among Older Adults: A Randomized Controlled Trial. Am J Med. 2015 Nov;128(11):1225-1236.e1. doi: 10.1016/j.amjmed.2015.06.017. Epub 2015 Jul 6. PMID: 26159634.

Ng TP, Ling LHA, Feng L, Nyunt MSZ, Feng L, Niti M, Tan BY, Chan G, Khoo SA, Chan SM, Yap P, Yap KB. Cognitive Effects of Multi-Domain Interventions Among Pre-Frail and Frail Community-Living Older Persons: Randomized Controlled Trial. J Gerontol A Biol Sci Med Sci. 2018 May 9;73(6):806-812. doi: 10.1093/gerona/glx207. PMID: 29069291.

Park JE, Jeon SY, Kim SA, Kim JH, Kim SH, Lee KW, Hwang YJ, Jung G, Suk HW, Park S, Lee DY. A Multidomain Intervention for Modifying Lifestyle Habits Reduces the Dementia Risk in Community-Dwelling Older Adults: A Single-Blinded Randomized Controlled Pilot Study. J Alzheimers Dis. 2019;70(1):51-60. doi: 10.3233/JAD-190016. PMID: 31127782.

Richard E, Moll van Charante EP, Hoevenaar-Blom MP, Coley N, Barbera M, van der Groep A, Meiller Y, Mangialasche F, Beishuizen CB, Jongstra S, van Middelaar T, Van Wanrooij LL, Ngandu T, Guillemont J, Andrieu S, Brayne C, Kivipelto M, Soininen H, Van Gool WA. Healthy ageing through internet counselling in the elderly (HATICE): a multinational, randomised controlled trial. Lancet Digit Health. 2019 Dec;1(8):e424-e434. doi: 10.1016/S2589-7500(19)30153-0. Epub 2019 Nov 14. PMID: 33323224.

Sipilä S, Tirkkonen A, Savikangas T, Hänninen T, Laukkanen P, Alen M, Fielding RA, Kivipelto M, Kulmala J, Rantanen T, Sihvonen SE, Sillanpää E, Stigsdotter Neely A, Törmäkangas T. Effects of physical and cognitive training on gait speed and cognition in older adults: A randomized controlled trial. Scand J Med Sci Sports. 2021 Jul;31(7):1518-1533. doi: 10.1111/sms.13960. Epub 2021 Apr 6. PMID: 33772877.

Sugimoto T, Araki A, Fujita H, Fujita K, Honda K, Inagaki N, Ishida T, Kato J, Kishi M, Kishino Y, Kobayashi K, Kouyama K, Kuroda Y, Kuwahata S, Matsumoto N, Murakami T, Noma H, Ogino J, Ogura M, Ohishi M, Shimada H, Sugimoto K, Takenaka T, Tamura Y, Tokuda H, Uchida K, Umegaki H, Sakurai T. Multidomain Intervention Trial for Preventing Cognitive Decline among Older Adults with Type 2 Diabetes: J-MIND-Diabetes. J Prev Alzheimers Dis. 2024;11(6):1604-1614. doi: 10.14283/jpad.2024.117. PMID: 39559873; PMCID: PMC11573805.

Tainta M, Ecay-Torres M, de Arriba M, Barandiaran M, Otaegui-Arrazola A, Iriondo A, Garcia-Sebastian M, Estanga A, Saldias J, Clerigue M, Gabilondo A, Ros N, Mugica J, Barandiaran A, Mangialasche F, Kivipelto M, Arrospide A, Mar J, Martinez-Lage P; GOIZ ZAINDU study group. GOIZ ZAINDU study: a FINGER-like multidomain lifestyle intervention feasibility randomized trial to prevent dementia in Southern Europe. Alzheimers Res Ther. 2024 Feb 27;16(1):44. doi: 10.1186/s13195-024-01393-z. PMID: 38413990; PMCID: PMC10898038.

Thunborg C, Wang R, Rosenberg A, Sindi S, Andersen P, Andrieu S, Broersen LM, Coley N, Couderc C, Duval CZ, Faxen-Irving G, Hagman G, Hallikainen M, Håkansson K, Kekkonen E, Lehtisalo J, Levak N, Mangialasche F, Pantel J, Rydström A, Stigsdotter-Neely A, Wimo A, Ngandu T, Soininen H, Hartmann T, Solomon A, Kivipelto M. Integrating a multimodal lifestyle intervention with medical food in prodromal Alzheimer's disease: the MIND-ADmini randomized controlled trial. Alzheimers Res Ther. 2024 May 30;16(1):118. doi: 10.1186/s13195-024-01468-x. PMID: 38812047; PMCID: PMC11138035.

Zülke AE, Pabst A, Luppa M, Roehr S, Seidling H, Oey A, Cardona MI, Blotenberg I, Bauer A, Weise S, Zöllinger I, Sanftenberg L, Brettschneider C, Döhring J, Lunden L, Czock D, Haefeli WE, Wiese B, Hoffmann W, Frese T, Gensichen J, König HH, Kaduszkiewicz H, Thyrian JR,

Riedel-Heller SG. A multidomain intervention against cognitive decline in an at-risk-population in Germany: Results from the cluster-randomized AgeWell.de trial. Alzheimers Dement. 2024 Jan;20(1):615-628. doi: 10.1002/alz.13486. Epub 2023 Sep 28. PMID: 37768074; PMCID: PMC10917033.

Xu X, Chew KA, Wong ZX, Phua AKS, Chong EJY, Teo CKL, Sathe N, Chooi YC, Chia WPF, Henry CJ, Chew E, Wang M, Maier AB, Kandiah N, Chen CL. The SINgapore GERiatric Intervention Study to Reduce Cognitive Decline and Physical Frailty (SINGER): Study Design and Protocol. J Prev Alzheimers Dis. 2022;9(1):40-48. doi: 10.14283/jpad.2022.5. PMID: 35098972; PMCID: PMC8753332.
