## Supplemental Figures for "Multidomain Therapy for Alzheimer’s Disease: A Scoping Review"

### Supplemental Figures
### VISUALIZATION OF TRIAL PARAMETERS

**Supplemental Figure 1**. Scatterplot matrix of values reported in **Table 2**. Overall, there is not much correlation between any trial parameters. However, trial size (N) and length of trial (Months) are negatively correlated with baseline FAST, suggesting that it is harder to recruit and maintain enrollment for individuals with more advanced Alzheimer's disease (AD). Larger trials tend to last longer. Observed effect size and significance are correlated, as expected from statistical principles. Trial size and observed effect size are anti-correlated, suggesting the possibility of a confounding factor (see Supplemental Figures 2-4). Date is the year of first participant enrollment.

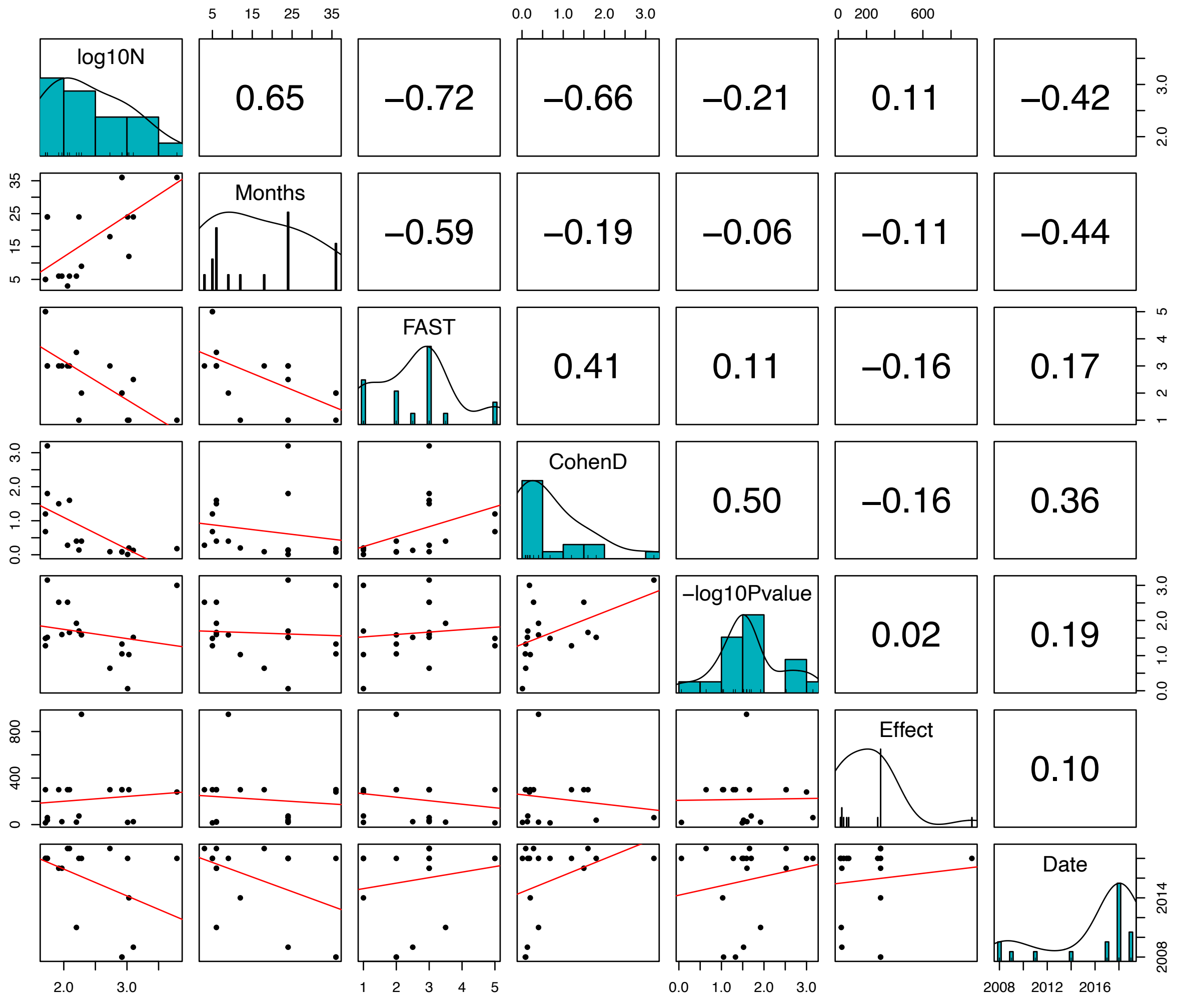

**Supplemental Figure 2**. Violin plot of the relationship between effect size and choosing either a composite Z-score or a validated single cognitive test for the primary trial outcome measure. Trials with simple measures tend to demonstrate greater effects.

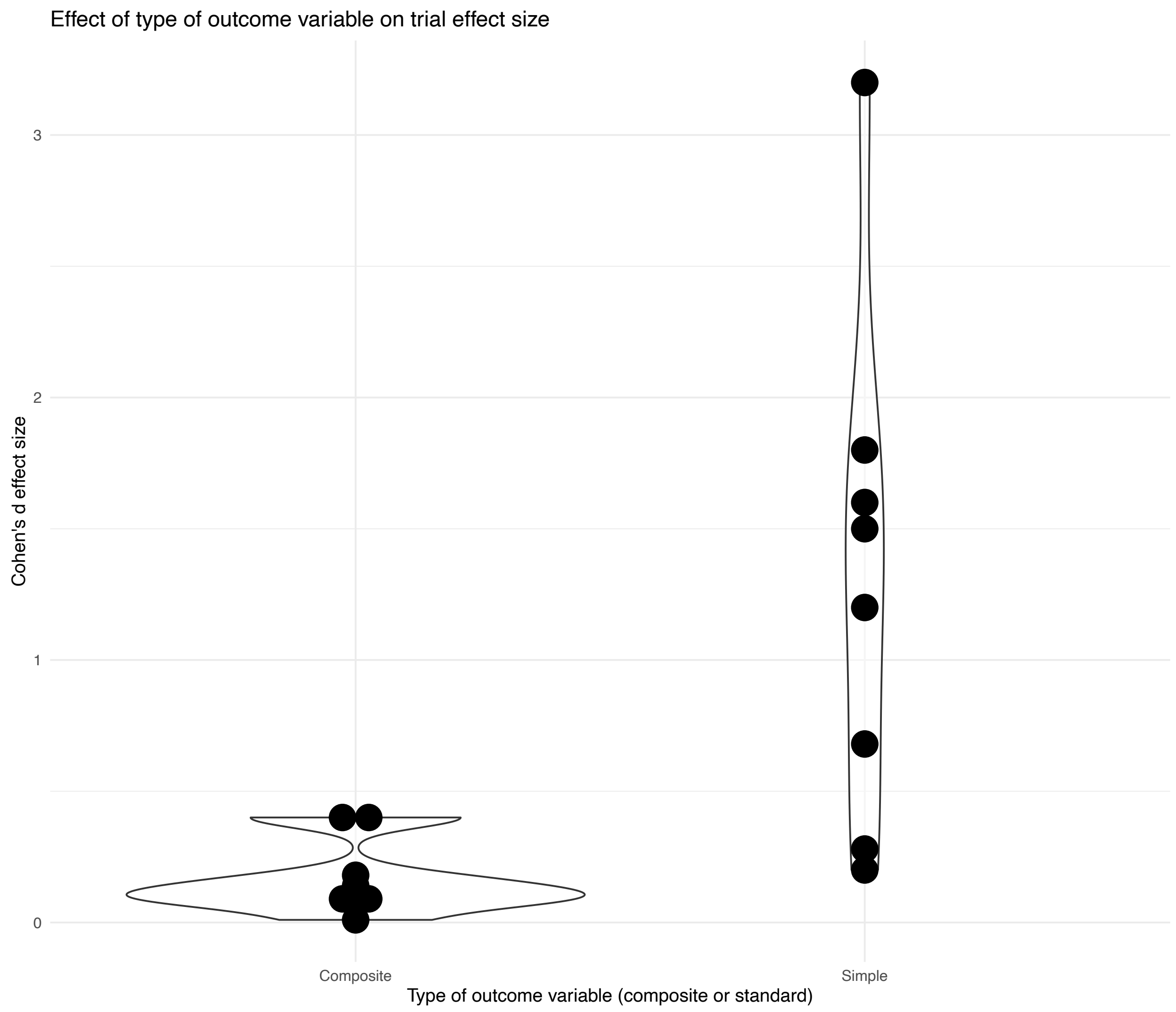

**Supplemental Figure 3**. Violin plot of the relationship between significance and choosing either a composite Z-score or a validated single cognitive test for the primary trial outcome measure. Trials with simple measures tend to demonstrate more significance (i.e., lower p-values).

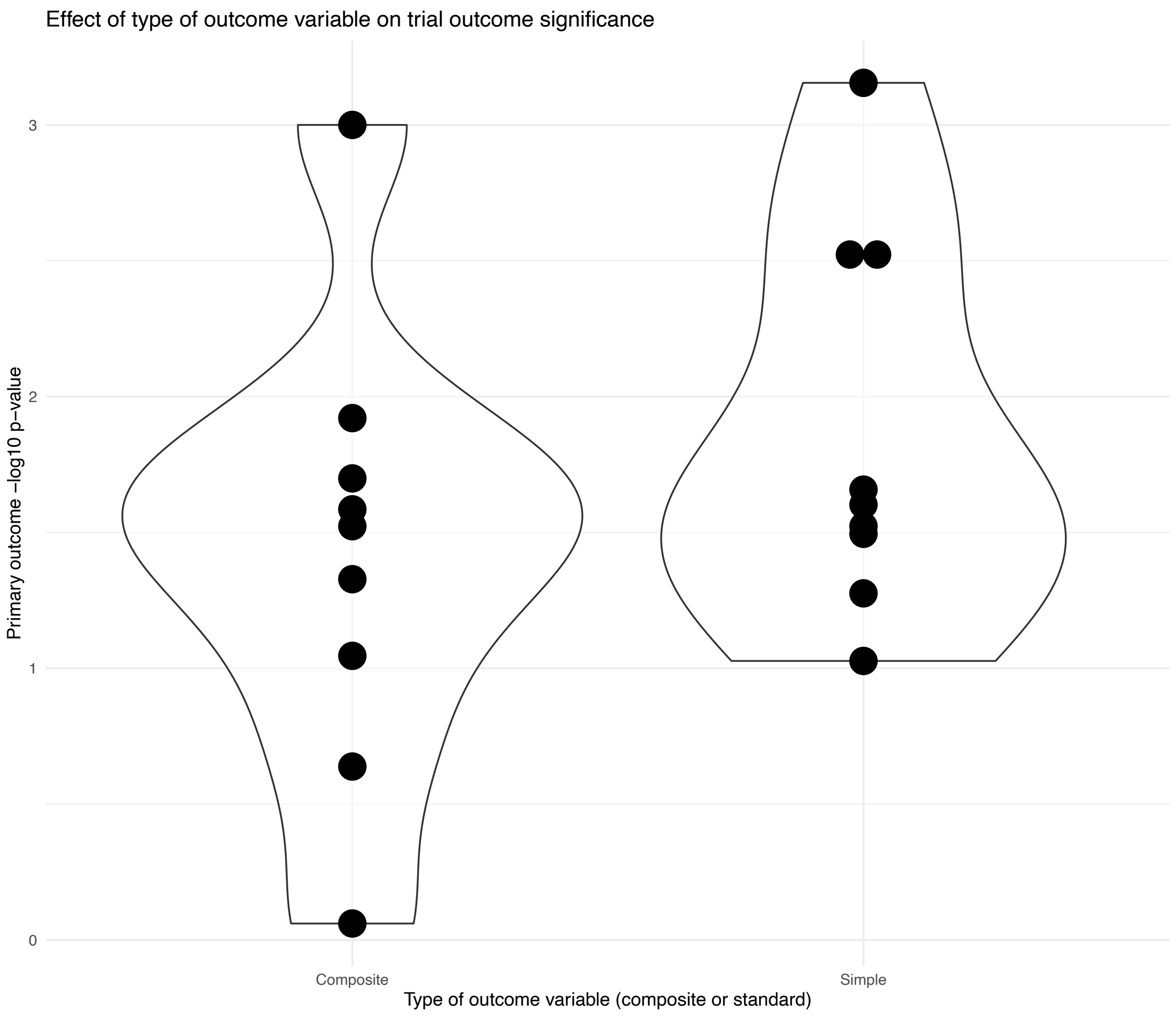

**Supplemental Figure 4.** Violin plot of the relationship between trial size and type of primary outcome variable. Larger trial designs tend to also be designed with composite Z-scores as their primary outcome measure.

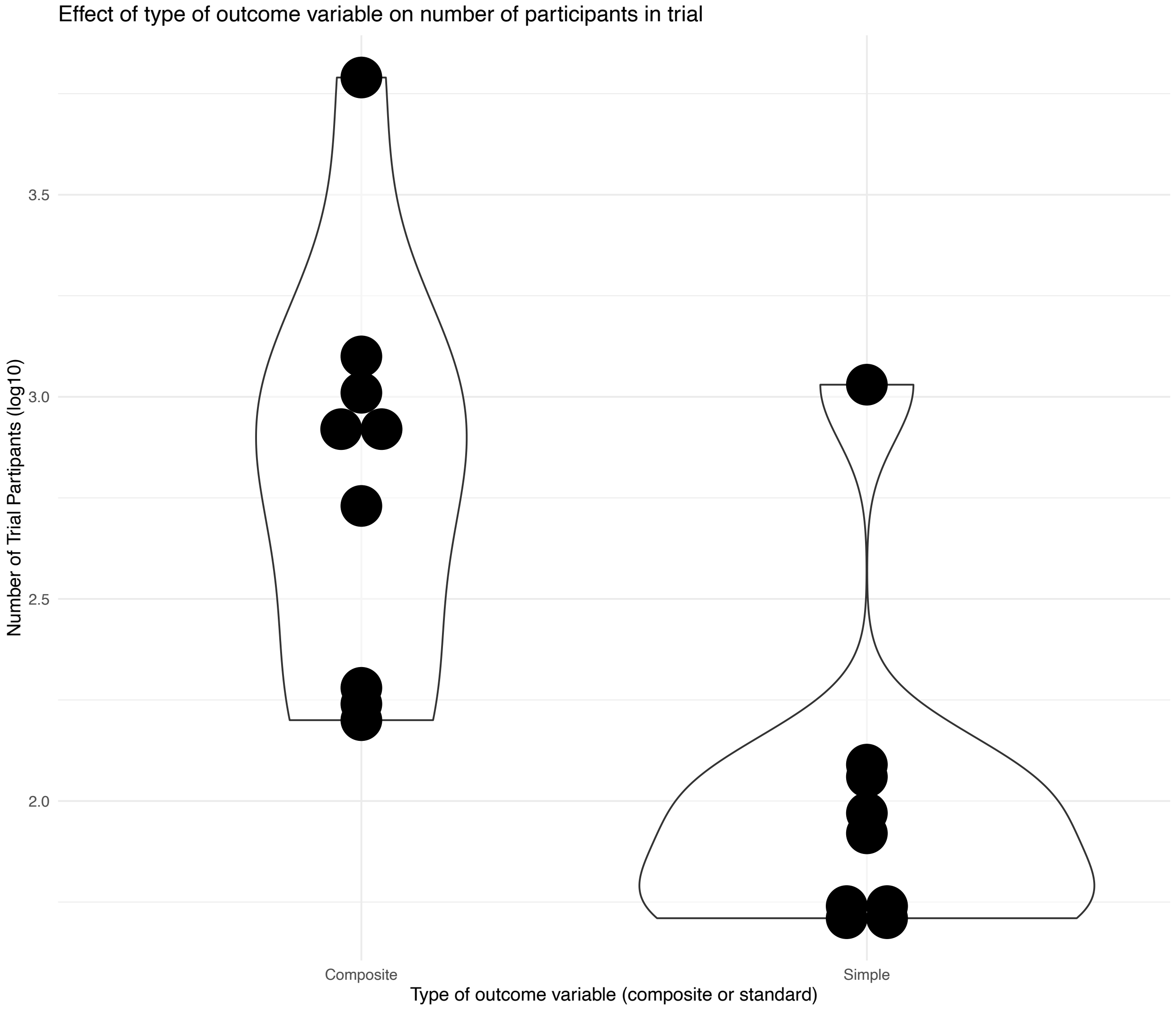

**Supplemental Figure 5**. Visualization of values reported in **Table 1**. Many factors affect the outcome in multidomain clinical studies. Important factors may include: (1) the number of domains of intervention, (2) the size of the study, (3) the age of participants at baseline, (4) the cognitive status or stage of disease at baseline, (5) the length of the study, and (6) the epoch and related environmental variables—such as COVID or current medical guidelines for managing comorbidities. Almost no pair of studies matches each other to a great extent for any of these parameters. Drawing conclusions from analyses or considerations of all studies together is therefore only possible for generating broad generalities. **(A) Scatterplot.** Area of each point ~ N; shading represents statistical significance (darker shades are more significant); vertical bars indicate age span at enrollment with the point placed at the average of the min and max; length of blue horizontal bars = length of the study; width of blue horizontal bars ~ effect size. **(B) Parallel coordinates plot.** Each variable is rescaled by subtracting the mean and dividing by the standard deviation.

**A.**

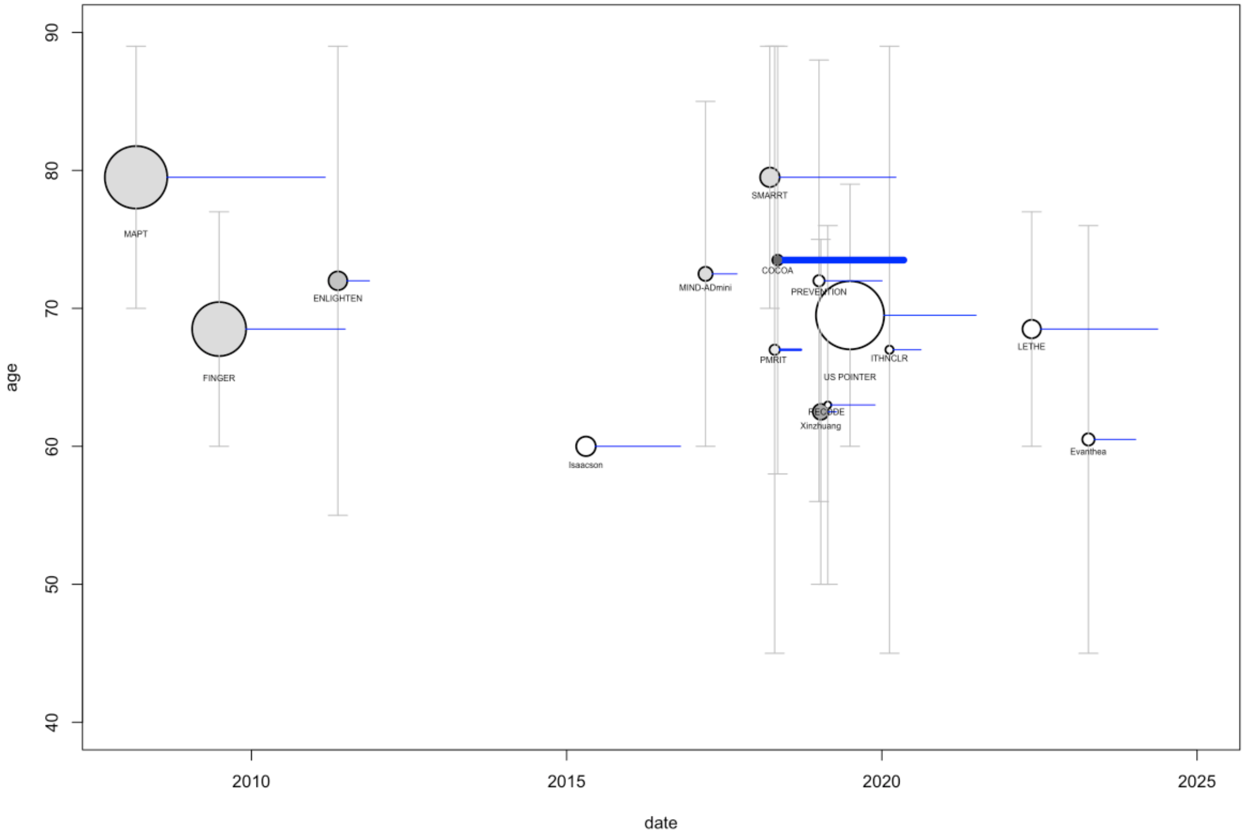

**B.**

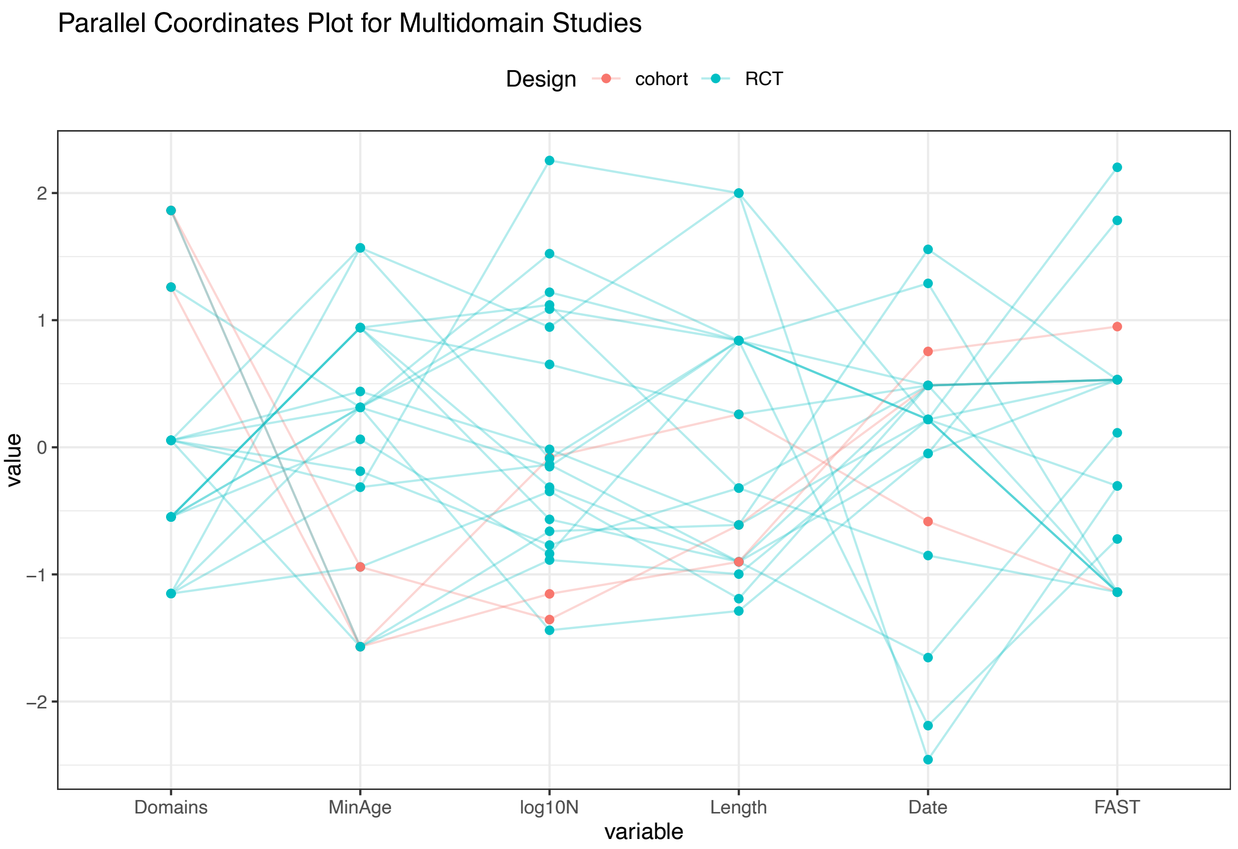
Parallel Coordinates Plot for Multidomain Studies
Design
cohort
RCT
value
2
1
0
−1
−2
Domains
MinAge
log10N
Length
Date
FAST
variable
